# Methylome-wide association study of antidepressant use in Generation Scotland and the Netherlands Twin Register implicates the innate immune system

**DOI:** 10.1101/2020.10.06.20207621

**Authors:** MC Barbu, F Huider, A Campbell, C Amador, MJ Adams, ME Lynall, DM Howard, RM Walker, SW Morris, J Van Dongen, DJ Porteous, KL Evans, E Bullmore, G Willemsen, DI Boomsma, HC Whalley, AM McIntosh

## Abstract

Antidepressants are an effective treatment for major depressive disorder (MDD), although individual response is unpredictable and highly variable. Whilst the mode of action of antidepressants is incompletely understood, many medications are associated with changes in DNA methylation in genes that are plausibly linked to their mechanisms. Studies of DNA methylation may therefore reveal the biological processes underpinning the efficacy and side effects of antidepressants.

We performed a methylome-wide association study (MWAS) of self-reported antidepressant use accounting for lifestyle factors and MDD in Generation Scotland (GS:SFHS, N=6,428, EPIC array) and the Netherlands Twin Register (NTR, N=2,449, 450K array) and ran a meta-analysis of antidepressant use across these two cohorts.

We found 10 CpG sites significantly associated with self-reported antidepressant use in GS:SFHS, with the top CpG located within a gene previously associated with mental health disorders, *ATP6V1B2* (β=-0.055, p_corrected_=0.005). Other top loci were annotated to genes including *CASP10, TMBIM1, MAPKAPK3, and HEBP2*, which have previously been implicated in the innate immune response. Next, using penalised regression, we trained a methylation-based score of self-reported antidepressant use in a subset of 3,799 GS:SFHS individuals that predicted antidepressant use in a second subset of GS:SFHS (N=3,360, β=0.377, p=3.12×10^−11^, R^2^=2.12%). In an MWAS analysis of prescribed selective serotonin reuptake inhibitors, we showed convergent findings with those based on self-report. In NTR, we did not find any CpGs significantly associated with antidepressant use. The meta-analysis identified the two CpGs of the ten above that were common to the two arrays used as being significantly associated with antidepressant use, although the effect was in the opposite direction for one of them.

Antidepressants were associated with epigenetic alterations in loci previously associated with mental health disorders and the innate immune system. These changes predicted self-reported antidepressant use in a subset of GS:SFHS and identified processes that may be relevant to our mechanistic understanding of clinically relevant antidepressant drug actions and side effects.

## Introduction

Major Depressive Disorder (MDD) is a leading cause of disability worldwide (1) and is caused by a combination of environmental and genetic factors (2). MDD has a number of effective treatments, with antidepressant drugs amongst the most commonly prescribed evidence-based therapies worldwide (3,4). Response to antidepressants is nevertheless unpredictable and highly variable, with only approximately 50% of individuals achieving remission after completing two treatments (5). The unmet needs of many individuals with MDD indicate the urgency of understanding the mechanisms of effective antidepressant action (6) so that their efficacy can be improved and their delivery targeted to those most likely to benefit.

The mode of action of antidepressants was originally assumed to be through the inhibition of monoamine re-uptake. However, since synaptic monoamine concentration poorly mirrors the trajectory of symptomatic improvement, their mechanism of action remains uncertain (7). Studies of antidepressants in both animal and human studies have implicated a number of possible modes of action. These include evidence that antidepressants lead to changes in DNA methylation (DNAm) and gene expression in a number of potentially relevant pathways (8). Francois et al. (2015) (9) demonstrated that antidepressants reversed stress-induced changes in DNAm and expression of 5*-HT1A* in an animal model, while Zimmermann et al. (2012) found that antidepressants affect activity of DNA methyltransferase 1 (*DNMT1*) (10). These studies demonstrate that antidepressants may lead to epigenetic changes that reveal their biologically relevant mechanisms of action.

Epigenetic processes are associated with alterations in DNA activity without alterations to the underlying genome sequence (11). DNAm is perhaps the most commonly investigated epigenetic change, owing to the availability of reliable high-throughput array technologies that can identify changes in DNAm at over 800K locations throughout the genome. Methylome-wide association studies (MWAS) have begun to identify a number of cytosine-phosphate-guanine (CpG) sites that are associated with MDD (12) and relevant environmental risk factors (13). DNAm changes may therefore capture an environmental archive of exogenous factors, including drug treatments, relevant to the onset and maintenance of MDD.

Recent studies have shown the potential of a DNAm-based risk score to predict MDD and its associated lifestyle and environmental factors, including antidepressant use (14,15). We previously (15) trained a DNAm predictor of MDD in 3,047 individuals using a penalised regression model and showed that it was significantly associated with self-reported antidepressant use in an independent cohort. This suggests that CpG sites conferring risk to MDD may also be linked to antidepressant use. A methylation-based predictor of self-reported antidepressant use may therefore be able to predict antidepressant use and its effects in other samples and may signpost the development of clinical biomarkers quantifying drug action.

Here, we sought to identify the DNAm changes associated with self-reported antidepressant use in 6,428 individuals (N_antidepressant use_=740) from the Generation Scotland study (GS:SFHS) using the Illumina Infinium MethylationEPIC array (16), capturing DNAm at approximately 850K sites and in 2,449 individuals (N_antidepressant use_=74) from the Netherlands Twin Register (NTR) using the Infinium HumanMethylation450 BeadChip Kit (17), capturing DNAm at 450K sites. We then conducted a meta-analysis using the two cohorts. Further, by splitting GS:SFHS into two DNAm datasets where the number of participants differed according to data availability, we were able to train a DNAm-based methylation score (MS) of self-reported antidepressant use in one dataset (N=3,799; N_antidepressant use_=585) and test its ability to predict self-reported antidepressant use in a second (N=3,360; N_antidepressant use_=317). We also addressed confounding by indication and by smoking in planned sensitivity analyses (18). Using linked National Health Service (NHS) Scotland medication prescribing data, we were able to identify whether the MWAS findings from self-report were also found when using contemporaneous electronic healthcare information collected around the time of the blood draw.

## Methods

### Study populations

#### Generation Scotland – the Scottish Family Health Study (GS:SFHS)

GS:SFHS is a family-based population cohort designed to investigate the genetic and environmental causes of common diseases and well-being in approximately 24,000 participants aged 18-98 years in Scotland. Baseline data was collected between 2006 and 2011 (19,20) and contains detailed information on a broad range of variables. DNA is also available from blood samples taken on more than 20,000 consenting participants.

GS:SFHS received ethical approval from NHS Tayside Research Ethics Committee (REC reference number 05/S1401/89) and has Research Tissue Bank Status (reference: 20/ES/0021). Written informed consent was obtained from all participants.

#### Netherlands Twin Register (NTR)

NTR is a population-based cohort of over 200,000 participants, consisting of twins, their parents, spouses, and siblings. Between 2004 and 2010, biosamples were collected in the NTR-Biobank project. Full details have been reported previously (21,22). DNA was collected from peripheral whole blood.

Informed consent was obtained from all participants. The study was approved by the Central Ethics Committee on Research Involving Human Subjects of the VU University Medical Centre, Amsterdam, an Institutional Review Board certified by the U.S. Office of Human Research Protections (IRB number IRB00002991 under Federal-wide Assurance-FWA00017598; IRB/institute codes, NTR 03-180).

### Phenotypes

#### GS:SFHS

##### Self-reported antidepressant use

For self-reported antidepressant use, a text-based questionnaire was sent to participants 1-2 weeks before attending an in-person appointment where DNAm was collected. The questionnaire recorded medication use through a “yes/no” checkbox with the following accompanying question: “Are you regularly taking any of the following medicationsã” where available options included “Antidepressants”.

Demographic data on individuals with self-reported antidepressant use is presented in Table 1. Participants with no self-reported antidepressant use were defined as those individuals who answered “No” to the “Antidepressants” sub-section of the questionnaire. There were 6,428 individuals with available self-reported data who also had DNAm, lifestyle, and MDD status data available.

**Table 1.** Demographic characteristics for GS individuals with self-reported antidepressant use and SSRI prescribing data in 12 months prior to blood draw date included in MWAS, including lifestyle variables and MDD; AD=antidepressant.

|  | Self-reported AD use sample |  | SSRI sample from NHS Scotland records |  |
| --- | --- | --- | --- | --- |
| Demographic characteristics | AD use (N=740) | No AD use (N=5,688) | SSRI use in 12-month interval (N=401) | No SSRI use (N=6,705) |
| **Age** |  |  |  |  |
| Mean (SD), range | 51.33 (11.07), 18 - 87 | 49.5 (13.73), 18 - 87 | 49.16 (12.35), 18 - 85 | 50.21 (13.65), 18 - 94 |
| **Sex (%)** |  |  |  |  |
| Female | 564 (76%) | 3,336 (59%) | 364 (75%) | 3,631 (54%) |
| Male | 176 (24%) | 2,352 (41%) | 123 (25%) | 3,074 (46%) |
| **Wave (%)** |  |  |  |  |
| 1 | 480 (65%) | 2,918 (51%) | 260 (65%) | 3,389 (51%) |
| 2 | 260 (35%) | 2,770 (49%) | 141 (35%) | 3,316 (49%) |
| **BMI** |  |  |  |  |
| Mean (SD), range | 28.17 (5.58), 16.11 - 51.29 | 26.60 (4.95), 14.78 - 67.62 | 28.37 (5.97), 17.50 - 51.29 | 26.68 (4.96), 15.93 - 67.62 |
| **Alcohol units** |  |  |  |  |
| Mean (SD), range | 8.99 (11.75), 0 - 105 | 10.56 (11.36), 0 - 146 | 9.46 (10.59), 0 - 72 | 10.91 (12.24), 0 - 326 |
| **Smoking status (%)** |  |  |  |  |
| Current smoker | 192 (26%) | 861 (15%) | 104 (26%) | 1,006 (15%) |
| Former smokers (quit < 1 year ago) | 15 (2%) | 146 (2%) | 2 (1%) | 154 (2%) |
| Former smokers (quit > 1 year ago) | 229 (31%) | 1,634 (29%) | 116 (30%) | 1,907 (29%) |
| Never smoked tobacco | 304 (41%) | 3,047 (54%) | 179 (43%) | 3,638 (54%) |
| **Pack years** |  |  |  |  |
| Mean (SD), range | 11.03 (16.03), 0 - 116 | 7.01 (13.51), 0 - 133 | 9.59 (14.58), 0 - 72.85 | 7.27 (14.07), 0 - 133 |
| **MDD status (%)** |  |  |  |  |
| Cases | 420 (57%) | 740 (13%) | 201 (50%) | 706 (11%) |
| Controls | 320 (43%) | 4,948 (87%) | 200 (50%) | 5,999 (89%) |

##### Selective serotonin reuptake inhibitors (SSRI) prescription and data linkage

Here, we selected antidepressants from BNF paragraph code 4.3.3 containing the following SSRIs: Citalopram, Escitalopram, Fluoxetine, Fluvoxamine Maleate, Paroxetine, and Sertraline (23) for individuals with medication data in GS:SFHS (see Supplementary Materials for more information). To investigate DNAm signatures of SSRI use, we restricted analyses to those individuals with SSRI dispensing records in the 12 months prior to blood draw date (N=401). Individuals with DNAm data that did not have any SSRI dispensed prescription records were marked as controls (N=5,688).

We used dates of dispensing, not prescription, when restricting antidepressant use based on time interval. Demographic data on individuals included here is presented in Table 1. Further details with regards to SSRI prescriptions are presented in Supplementary Table 1.

##### Lifestyle factors and MDD status

Body mass index (BMI) was computed using height (cm) and weight (kg) as measured by clinical staff at baseline recruitment. Participants reported the number of units of alcohol consumed during the past week and their smoking status (never, former, current); pack years was used to measure heaviness of smoking in current smokers (24).

MDD status was assessed using the Structured Clinical Interview of the Diagnostic and Statistical Manual, version IV (SCID) (25). Participants with no MDD were defined as those individuals who did not fulfil criteria for a current or previous MDD diagnosis following the SCID interview (26).

Further details about other diagnoses and medication intake are presented in Supplementary Tables 2 and 3. Participants who suffered from other psychiatric disorders (e.g. bipolar disorder, psychosis) were excluded from analyses (N=11).

#### NTR

Demographic data on individuals with antidepressant use is presented in Table 2.

##### Self-reported antidepressant use

At blood draw, participants were asked about all current medication use and for all medicines the dose, brand and chemical names were recorded directly from the medication packaging. Following the Anatomical Therapeutic Chemical (ATC) classification system, subclasses of N06A were considered antidepressants. Antidepressant medication use was coded as 1 (current user) / 0 (non-user).

**Table 2.** Demographic characteristics for NTR individuals with self-reported antidepressant use data within a week of blood draw included in MWAS, including lifestyle variables and MDD; AD=antidepressant.

| Demographic characteristics | Self-reported AD use sample |  |
| --- | --- | --- |
|  | AD use<br>(N=74) | No AD use<br>(N=2,375) |
| **Age** |  |  |
| Mean (SD), range | 37.74 (12.20),<br>18 – 72 | 36.76 (3.97),<br>18 - 80 |
| **Sex (%)** |  |  |
| Female | 52 (70%) | 1,618 (68%) |
| Male | 22 (30%) | 757 (32%) |
| **BMI** |  |  |
| Mean (SD), range | 25.06 (4.25), 18.30 – 34.60 | 24.24 (3.97), 14.50 – 50.70 |
| **Smoking status (%)** |  |  |
| Current smokers | 13 (17%) | 434 (18%) |
| Former smokers | 19 (26%) | 506 (21%) |
| Never smoked tobacco | 42 (57%) | 1,435 (61%) |
| **Pack years** |  |  |
| Mean (SD), range | 3.56 (5.83),<br>0 - 25 | 3.95 (8.69),<br>0 – 105 |
| **MDD status (%)** |  |  |
| Cases | 10 (14%) | 395 (17%) |
| Controls | 64 (86%) | 1,980 (83%) |

##### Lifestyle factors and MDD status

Body mass index (kg/m2) was computed based on weight and height obtained at the time of blood sampling. Current and past smoking behaviour was dummy-coded as current/former/never smokers and pack-years were recorded as the number of cigarettes smoked per day/20⍰× ⍰number of years smoked.

Lifetime MDD was derived from the Composite International Diagnostic Interview (CIDI) (27), the Lifetime Depression Assessment Self-report (LIDAS) (28), and DSM-oriented Adult Self Report scales (ASR) of the Achenbach System of Empirically Based Assessment (ASEBA) (29) (Supplementary Materials). Data from the CIDI, LIDAS and ASR scales were combined to form one numeric variable coded as 1 (lifetime MDD case) / 0 (lifetime MDD control). When data from multiple sources were available for an individual, the information from CIDI was favoured over that of LIDAS and ASR, and data from LIDAS over that of ASR.

#### DNA methylation

##### GS:SFHS

Genome-wide DNAm data profiled from whole blood samples was available for 9,873 individuals in GS:SFHS using the Illumina Human-MethylationEPIC BeadChip (16). DNAm data for individuals was initially released in two waves (wave 1_N_=5101; wave 2_N_=4,450). Quality control (QC) and normalisation were conducted using R packages ShinyMethyl (30) and watermelon (31). Details of the protocol are described in the Supplementary Materials.

##### NTR

DNAm in the NTR-Biobank study (21,32,33) was assessed with the Infinium HumanMethylation450 BeadChip Kit ((17), Illumina, San Diego, CA, USA) by the Human Genotyping facility (HugeF) of ErasmusMC, the Netherlands (http://www.glimdna.org/) as part of the Biobank-based Integrative Omics Study (BIOS) consortium (34). DNAm measurements have been described previously (33,34) (see Supplementary Materials for more information).

### Statistical methods

#### Methylome-wide association

##### GS:SFHS

We used linear regression models run in the “limma” package (35) in R to analyse the association of each CpG site, included as an outcome variable, with self-reported antidepressant use included as the predictor variable. The R code for the current analyses is available in the Supplementary Materials. The following covariates were included: age, sex, wave (indicating different DNAm data pre-processing waves), as well as BMI, alcohol units, smoking status, and pack years to observe whether the inclusion of lifestyle factors attenuates the effect of self-reported antidepressant use. MDD status was also included to observe whether associations between CpGs and self-reported antidepressant use were influenced by the presence of MDD and potentially confounded by the indication for prescription, as some individuals not classed as having MDD have indicated antidepressant use (N=320). There were 674,246 CpGs available after QC and a Bonferroni correction (0.05/674,246) was used to define methylome-wide significance (p≤ 7.42×10^−8^).

Using the same model as above, we further carried out sensitivity analyses in (1) non-smokers (N=3,351, N_antidepressant use_=304), (2) individuals with (N=5,368, antidepressant users=420) and without (N=5,268, antidepressant users=320) MDD, and (3) individuals with SSRI dispense dates within 12 months prior to blood draw date (N=7,106, N_antidepressant use_=401) (see Supplementary Materials for more information).

We compared findings in the main antidepressant use MWAS with summary statistics from a MDD MWAS in the same sample (N=8,844, MDD cases=1,461), by looking at effect sizes for significant CpGs in both analyses (Supplementary Figures 2 and 3).

##### NTR

The association between antidepressant medication and DNAm was tested under a generalized estimation equation (GEE) model with DNAm M-values as outcome and self-reported antidepressant use as a predictor. The model included the following covariates: MDD status, sex, age at blood sampling, percentages of monocytes, eosinophils, and neutrophils, HM450k array row, 96-wells bisulfite sample plate (dummy coding), BMI, smoking, and smoking pack-years. MWAS analyses were performed with the R package ‘gee’. The following settings were used: Gaussian link function (for continuous data), 100 iterations, and the ‘exchangeable’ option to account for the correlation structure within families and within persons.

##### Meta-analysis

There were N=6,428 and N=2,449 participants in GS and NTR, respectively, and N=318,856 overlapping CpGs between the two cohorts. Meta-analysis of these two datasets was performed in METAL (36) using p-value based analysis (N=8,877). The meta-analysis was based on N=318,856 total overlapping CpGs across GS:SFHS and NTR and a Bonferroni correction (0.05/318,856) was used to define methylome-wide significance (p≤ 1.57×10^−7^).

##### DNAm score analysis

To create a training and testing dataset, we separated individuals in GS:SFHS by DNAm pre-processing wave (N_wave 1_=3,799; N_wave 2_=3,360). This number differed from the final number included in the MWAS (N=6,428) due to data availability for covariates included in different analyses. R package “biglasso” was used to train DNAm predictors (Supplementary Materials).

As in the MWAS, we carried out sensitivity analyses by training a score in wave 1 in (1) non-smokers (N=1,952, N_antidepressant use_=226) and (2) individuals with no MDD diagnosis (N=2,791, antidepressant users=195) (see Supplementary Materials).

There were 76, 20, and 35 CpG sites identified for the full (MS), smoker-excluded (MS-ns), and MDD-excluded (MS-control) datasets described above, respectively. The list of CpG sites and their corresponding weights are presented in Supplementary Tables 4, 5, and 6. MS, MS-ns, and MS-control were created for each individual in wave 2 (N=3,360; antidepressant users=317) by taking the sum of the product of the identified DNAm residualised M-values and supplied model coefficient values. Regression models were then run to identify associations between the scores and self-reported antidepressant use, lifestyle factors and MDD.

##### Pathway and regulatory element overlap analysis

We used the Infinium MethylationEPIC BeadChip database to annotate significant CpG sites in GS:SFHS to genes (16). The database provides information with regards to genes, chromosome location, start and end sites, and other features.

To assess pathway enrichment for differentially methylated CpG sites while correcting for biases in the representation of genes on the Infinium BeadChip, we used missMethyl (37), accessed via methylGSA (38). Gene Ontology (GO) terms were accessed using the msigdbr package (39). Pathways included in the analysis were all GO Biological Process pathways of size 20-500 genes inclusive. CpG sites included in the analysis were those significant at a threshold of p<1×10^−5^ (N=144), as used in previous studies (40).

To assess overlap of differentially methylated CpG sites with the 15 chromatin states across 127 tissues from the Consolidated Roadmap Epigenomics dataset, we used eFORGE 2.0 (41), accessed via the web tool at https://eforge.altiusinstitute.org and using default settings.

##### Identification of differentially-methylated regions (DMR)

We used the dmrff function implemented in the dmrff R package (42) to identify differentially methylated regions in GS:SFHS, defined as regions containing 2-30 CpG sites separated by ≤ 500 bp. DMR analysis uses CpGs with unadjusted MWAS p-values ≤ 0.05 and methylation changes in a consistent direction, and DMRs with a Bonferroni-corrected p-value ≤ 0.05 are considered statistically significant.

## Results

### Demographic characteristics

Individuals with self-reported antidepressant use who had incomplete lifestyle (BMI, alcohol consumption, and smoking) and disorder (MDD status) data were excluded. There were 6,428 individuals in the final MWAS of self-reported antidepressant use in GS:SFHS (N_antidepressant use_: 740) and 2,449 (N_antidepressant use_=74) in NTR. Descriptive and demographic characteristics for these individuals, as well as individuals with prescribing data, are presented in Table 1 for GS:SFHS and Table 2 for NTR.

### Methylome-wide association

#### GS:SFHS

##### Self-reported antidepressant use

MWAS identified 10 CpG sites that were associated with self-reported antidepressant use (p≤ 7.42×10^−8^). Nine CpG sites were significantly hypomethylated in antidepressant users, while 1 site (cg26277237) was hypermethylated. Information about each CpG site is shown in Table 3 and depicted as a Manhattan plot in Figure 1A.

**Table 3.**
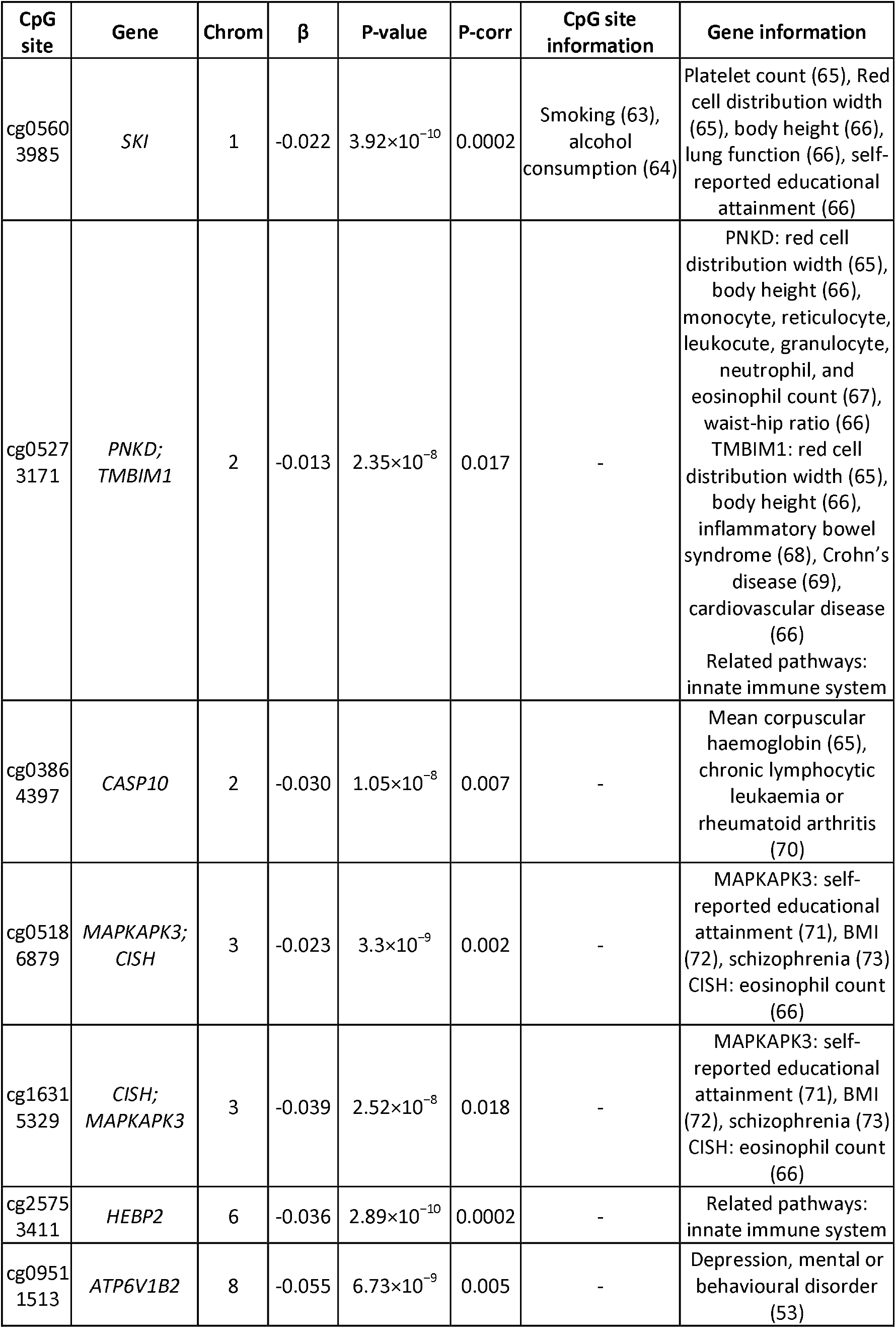
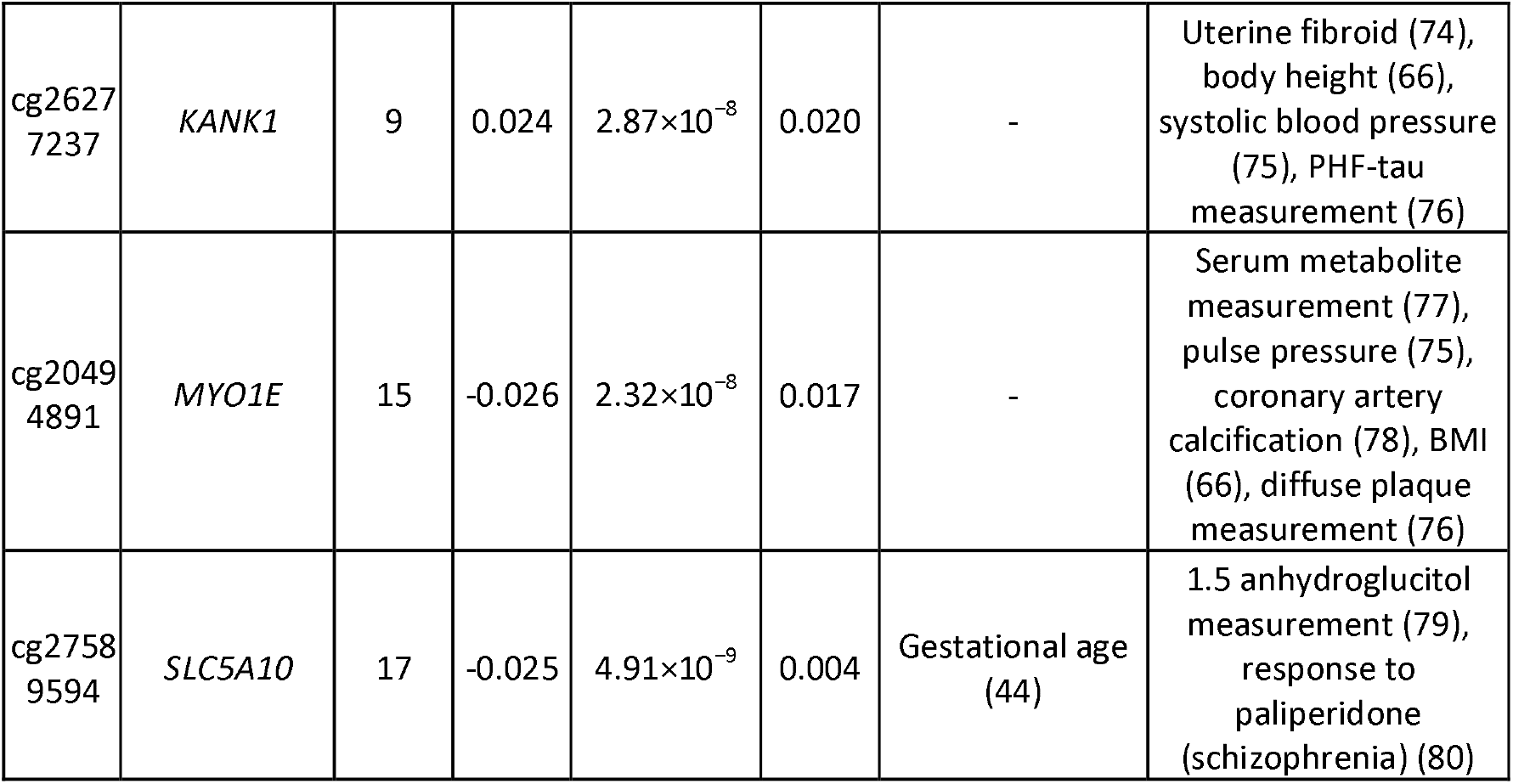
CpGs significantly associated with self-reported antidepressant use in GS (N=6,428; antidepressant use=740; N CpGs=10) along with gene annotations, chromosome, standardised effect size, nominal and Bonferroni-corrected p-values. Background information for each CpG and gene was extracted from EWAS (http://www.ewascatalog.org/; association between traits and CpGs on Illumina 450K array at p≤ 1.0×10^−4)^; and GWAS (https://www.ebi.ac.uk/gwas/; associations between traits and SNPs at p≤ 1.0×10^−5^) catalogue databases. All associations included in the table from these two catalogues are genome-wide significant.

**Figure 1A, 1B, 1C.**
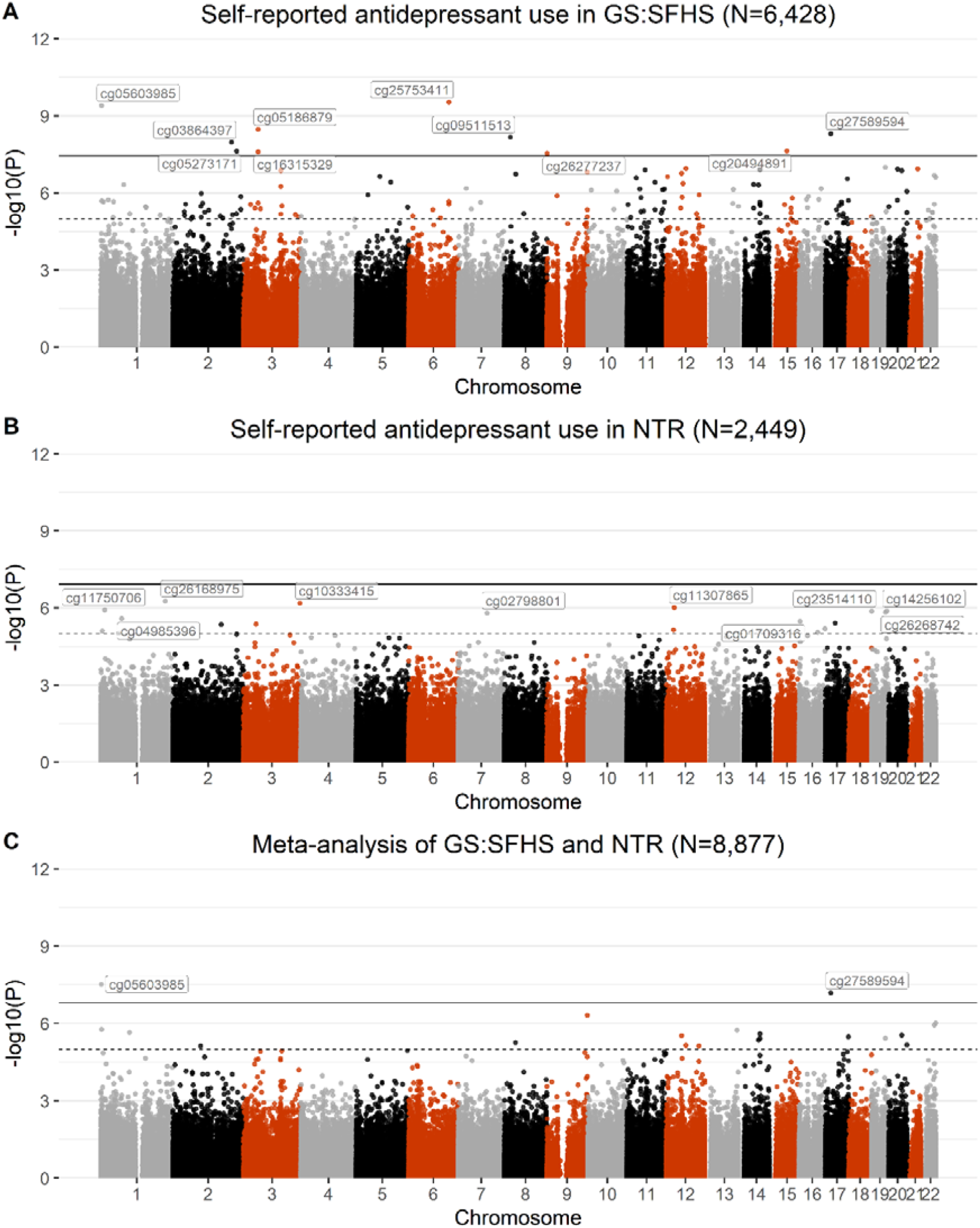
Manhattan plots showing MWAS of self-reported antidepressant use in GS:SFHS (1A), MWAS of self-reported antidepressant use in NTR (1B), and meta-analysis of MWAS in GS:SFHS and NTR (1C). The black line defines methylome-wide significance for each analysis (GS:SFHS: p≤ 3.6×10^−8^; NTR: p≤ 1.22×10^−7^; meta-analysis: p≤ 1.57×10^−8^) and the dotted line defines p≤ 1×10^−5^. Methylome-wide significant hits (GS:SFHS: 10; meta-analysis: 2) and the top 10 CpGs in NTR are labelled on the graph.

The EWAS catalogue (http://www.ewascatalog.org/) includes CpG sites represented on the Illumina 450K methylation array that have shown associations with traits at p≤ 1×10^−4^ in genome-wide analyses. The catalogue was used to cross-reference all CpG sites associated with self-reported antidepressant use with the wider literature. cg05603985 was previously found to be associated with smoking (p=1.8×10^−43^; (18)) and alcohol consumption (p=7.9×10^−13^ in African ancestry and p=1.9×10^−8^ in European ancestry; (43); cg27589594 was found to be associated with gestational age (p=3.5×10^−16^; (44)). Eight of the CpG sites were represented only on the Infinium MethylationEPIC BeadChip (containing approximately 850K CpG sites) and were therefore not included in the catalogue. Searches conducted on other databases, including EWASdb (45) indicate that these 8 CpG sites have not been previously associated with any trait.

The Pearson’s correlation between the MWAS effect sizes from the full-sample MWAS and MWAS in non-smokers across all CpGs was r=0.655 (95% C.I.=0.636-0.674). When restricting CpGs to those that reached a p-value≤ 5×10^−5^, the correlation increased further to r=0.877 (95% C.I.=0.869-0.884), and Bonferroni-corrected CpGs (N=10) in the full sample and the same CpGs in the non-smoker sample had a high correlation (r=0.980, 95% C.I.=0.979-0.981). Supplementary Table 7 indicates the effect size and nominal p-value for all CpGs found to be associated with self-reported antidepressant use in the full sample and in the non-smoker sample.

The Pearson’s correlation between effect sizes obtained from the MWAS of antidepressant use in individuals with MDD and MWAS of antidepressant use in those without MDD across all CpGs was *r*=0.094 (95% C.I.=0.069-0.118). Restricting CpGs to a significance cut-off of p≤ 5×10^−5^ increased the correlation in effect sizes to *r*=0.961 (95% C.I.=0.959-0.963). Supplementary Table 8 indicates the effect size and nominal p-value for all CpGs found to be associated with self-reported antidepressant use in the full sample and in the sensitivity analysis sample.

##### SSRI

MWAS conducted using individuals with dispensing data identified 12 CpG sites, all hypomethylated, associated with SSRI use (the most commonly prescribed treatment for depression) within 12 months prior to blood draw date, which are shown in Supplementary Table 9 and Supplementary Figure 1.

Additionally, all 10 CpG sites that were associated with self-reported antidepressant use (p≤ 7.42×10^−8^) were found to be nominally significant (p<0.05) in the MWAS including SSRI dispensing records as a phenotype. The direction of effect was consistent for all CpG sites identified in the self-report MWAS: 9 were hypomethylated and 1 was hypermethylated, presented in Supplementary Figure 1 and Supplementary Table 10 (β and p-value for smallest and largest associations included cg09511513 (β=-0.026, p=0.028) and cg26277237 (β=0.027, p=3.09×10^−7^)). The majority (10/12) of CpG sites identified in the MWAS of SSRI also were nominally significant in the MWAS where the self-report variable was input as the predictor (Supplementary Table 11).

##### NTR

There were no methylome-wide significant findings in NTR. The top ten CpG sites were annotated to genes implicated in psychiatric- and cognition-related phenotypes and in disorders of the immune system and are presented Supplementary Table 12. Figure 1B represents a Manhattan plot of NTR results.

##### Meta-analysis

Meta-analysis of antidepressant use MWAS across GS:SFHS and NTR revealed two Bonferroni-corrected CpG sites, which were on the 450K array; both were identified in the GS:SFHS MWAS; cg05603985 (β=-0.021, p=3.01×10^−8^, q=0.009) had an opposite direction of effect across cohorts (negative in GS:SFHS and positive in NTR), and cg27589594 (β=-0.024, p=6.46×10^−8^, q=0.021) had the same direction of effect (negative) in both. Methylome-wide results are depicted in Figure 1C.

There were N=318,856 overlapping CpGs between GS:SFHS and NTR, and effect sizes were not strongly correlated (Pearson’s correlation r=0.081, 95% C.I.=0.06-0.102). When restricting CpGs to those that reached a p-value≤ 0.05 across both MWAS (N=1,714), the correlation between effect sizes increased (r=0.32, 95% C.I.=0.301-0.339). Further information with regards to concordant signals across the two cohorts is in the Supplementary Materials.

##### DNAm score analysis

LASSO regression selected 76, 20, and 35 CpGs to calculate MS, MS-ns, and MS-control for self-reported antidepressant use, respectively, in GS:SFHS wave 2.

Briefly, the MS was associated with self-reported antidepressant use in a model adjusted for age, sex, and 10 genetic PCs (β=0.377, p=3.12×10^−11^, R^2^=2.12%); After adjustment for BMI, alcohol units, smoking status, pack years, and MDD, this association remained significant, although the variance explained decreased (β=0.213, p=0.0035, R^2^=0.56%). The association between MS-ns and self-reported antidepressant use was in the same direction but became non-significant, both in the covariate-free model (β=0.106, p=0.075, R^2^=0.16%) and in the model with the above covariates included (β= 0.04, p=0.565, R^2^=0.02%). Finally, MS-control was significantly associated with self-reported antidepressant use in the covariate-free model (β=0.254, p=1.15×10^−5^, R^2^=0.93%), but not when including covariates (β=0.064, p=0.381, R^2^=0.05%). Table 4 indicates associations with lifestyle traits and MDD for all scores.

**Table 4.** Association between MS (methylation score), MS-ns (methylation score trained on non-smokers), and MS-control (methylation score trained on individuals with no MDD diagnosis), and self-reported antidepressant use, MDD, and 4 lifestyle factors (BMI, smoking status, pack years, alcohol units). All regression models include age, sex, and 10 genetic principal components as covariates, except for “AD use*”, which also includes BMI, smoking status, pack years, alcohol units, and MDD as covariates. Effect sizes represent standardized betas. R^2^ represents the variance explained in the outcome variables by each score. The number of individuals in each model varies based on different available data for each variable.

| Outcome variable (N) | MS |  |  | MS-ns |  |  | MS-control |  |  |
| --- | --- | --- | --- | --- | --- | --- | --- | --- | --- |
| | $\beta$ | p-value | R<sup>2</sup> | $\beta$ | p-value | R<sup>2</sup> | $\beta$ | p-value | R<sup>2</sup> |
| AD use (3,360) | 0.377 | $3.12 \times 10^{-11}$ | 2.12% | 0.106 | 0.075 | 0.16% | 0.254 | $1.15 \times 10^{-5}$ | 0.93% |
| AD use* (3,009) | 0.213 | 0.0035 | 0.56% | 0.04 | 0.565 | 0.02% | 0.064 | 0.381 | 0.05% |
| MDD (3,360) | 0.28 | $5.78 \times 10^{-7}$ | 1.14% | 0.158 | 0.007 | 0.35% | 0.202 | 0.0004 | 0.57% |
| BMI (3336) | 0.088 | $2.54 \times 10^{-7}$ | 0.75% | 0.021 | 0.23 | 0.01% | 0.075 | $1.33 \times 10^{-5}$ | 0.53% |
| Alcohol units (3099) | 0.141 | $< 2 \times 10^{-16}$ | 1.93% | 0.007 | 0.665 | 0% | 0.127 | $5.08 \times 10^{-14}$ | 1.56% |
| Smoking status (3,322) | 0.632 | $< 2 \times 10^{-16}$ | 6.11% | 0.077 | 0.028 | 0.11% | 0.567 | $< 2 \times 10^{-16}$ | 5.08% |
| Pack years (3,310) | 0.305 | $< 2 \times 10^{-16}$ | 9.14% | 0.061 | 0.0004 | 0.34% | 0.287 | $< 2 \times 10^{-16}$ | 8.09% |

##### Pathway and regulatory element overlap analysis

Following FDR adjustment for multiple comparisons, the only over-represented pathways were related to regulation of myeloid cell differentiation (GO Negative Regulation of Myeloid Cell Differentiation and GO Negative Regulation of Myeloid Leucocyte Differentiation, p_adjusted_=0.04 for both). Enrichment of these pathways was driven by hypomethylation in the antidepressant use group in the body or 5’UTR of the following genes: *GAT2, HOXA7, INPP5D, MEIS1, RAR and UBASH3B*.

Testing for overlap of differentially methylated CpG sites at p<1×10^−5^ with chromatin states in 127 tissues from the Roadmap Epigenomics Consortium, the greatest enrichment was seen in blood cell tissues. Of the peripheral blood cell subsets tested, the cell type showing the greatest enrichment was monocytes (Figure 2), with significant enrichment for monocyte enhancers (q=4×10^−11^) and regions flanking active transcription start sites in monocytes (q=1×10^−9^).

**Figure 2.**
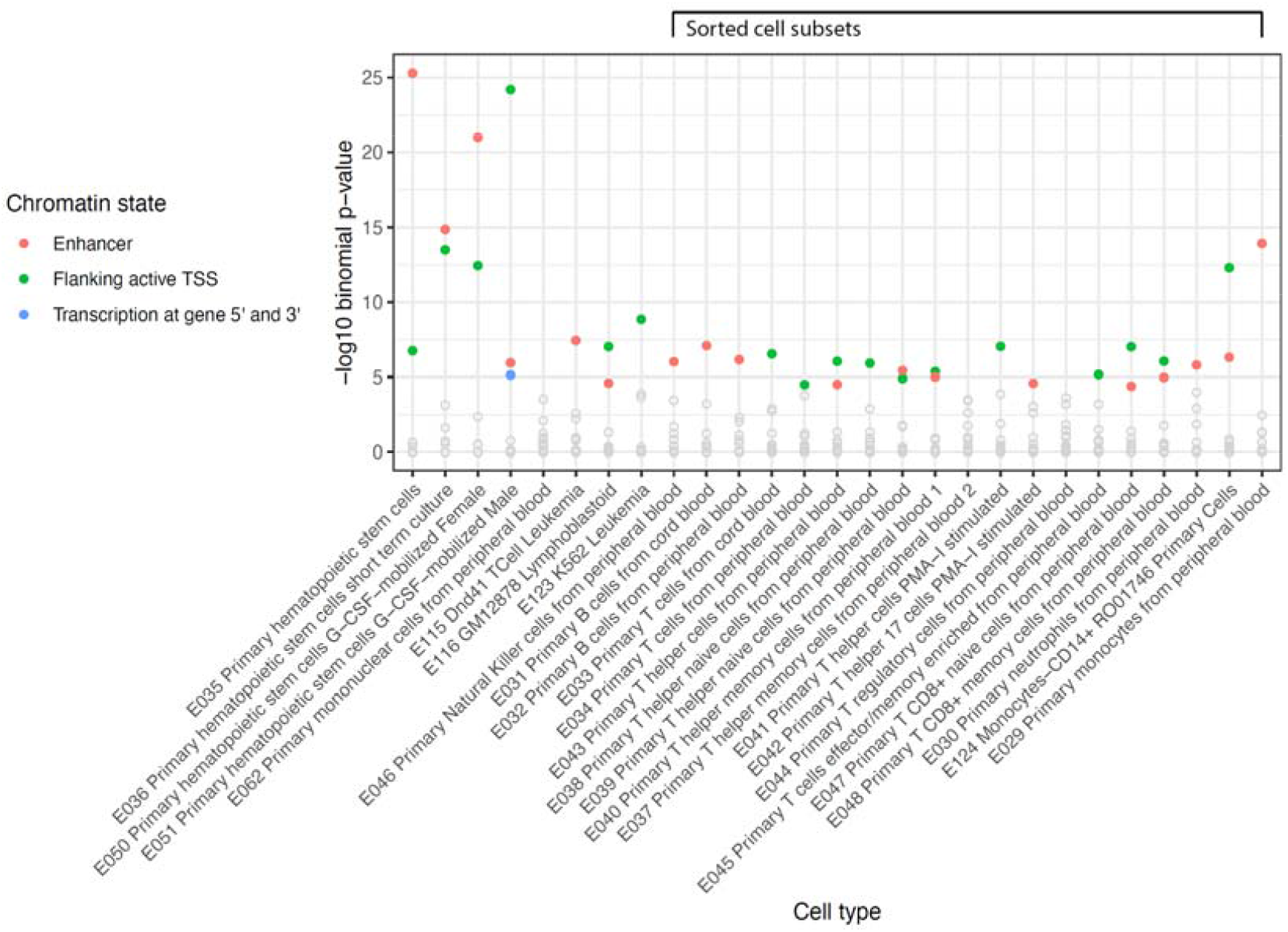
Differentially methylated CpG sites at p<1×10^−5^ in GS:SFHS enriched for peripheral blood cell subsets. The x-axis represents peripheral blood cell types, while the y-axis indicates corrected p-value.

We repeated the tests above using the results from the MEWAS of antidepressant prescriptions (N CpGs at p<1×10^−5^=484). We did not find enrichment of any specific GO biological processes but replicated the finding from the self-reported antidepressant use MWAS that monocytes were the peripheral blood cell subset most enriched in the MWAS signal (enhancers in primary monocytes from peripheral blood were enriched at q=4×10^−94^) and also found strong enrichment for enhancers in peripheral blood neutrophils (q=4×10^−79^).

##### DMR analysis

We identified 159 DMRs, ranging from 2-11 CpGs in length, to be associated with self-reported antidepressant use using the DMR adjusted p-values (Supplementary Excel File 1). The region with the largest significance (p.adjust=7.54×10^−12^) spanned 4 CpGs annotated to GATA2 on chromosome 3, a gene involved in stem cell maintenance and in which mutations are associated with wide-ranging immunological disorders (46,47). The largest DMR (p.adjust=2.04×10^−8^) spanned 11 CpGs located on chromosome 2, where most were annotated to OTX1, a gene previously associated with risk-taking behaviour (48). The gene may also play a role in the brain and development of sensory organs (49). Only one DMR spanning 2 CpGs (p.adjust=2.55×10^−7^) contained a CpG that was also associated with the trait at the level of individual probes (cg05186879, p=4.91×10^−9^).

## Discussion

Self-reported antidepressant use is associated with differences in DNAm at 10 CpG sites in GS:SFHS located in genes that have previously been associated with psychiatric disorders and the innate immune system. There were no methylome-wide findings in NTR. We found that all 10 CpG sites in GS:SFHS were also nominally associated with antidepressant prescription data obtained from the electronic health record, indicating agreement between self-reported and record linkage data. Sensitivity analyses in GS:SFHS indicated that results were not significantly confounded by smoking. As shown in Supplementary Table 7, the effect for all 10 associated CpGs are in the same direction across sensitivity analyses, and all p-values in the sensitivity analyses are nominally significant. The effect sizes of the top 10 CpGs were also highly correlated in the full sample and in the non-smoker sample (*r*=0.980). A DNAm score trained on one sub-sample was associated with self-reported antidepressant use, four lifestyle factors, and MDD in a second unrelated sample from the same study. Pathway analysis revealed an enrichment of differentially methylated genes in myeloid cell differentiation. However, the effect size correlation between the 10 associated CpGs and the same CpGs in a MDD MWAS (r=0.674) may indicate that DNAm signatures of antidepressants act as a proxy for MDD, although further analyses with much larger samples will be needed to disentangle the effects between antidepressant use and MDD on DNAm. Lastly, meta-analysis of MWAS in GS:SFHS and NTR identified two CpGs that were also found in GS:SFHS.

The CpG site with the largest effect size, both in GS:SFHS and the meta-analysis (β=-0.055) was cg09511513, located in *ATP6V1B2*. This CpG site has not been associated with any other traits previously, to the best of our knowledge. *ATP6V1B2* encodes a component of vacuolar ATPase, which is a multisubunit enzyme that mediates the acidification of eukaryotic intracellular organelles, including endosomes and lysosomes, and may be involved in neurotransmission (50). Importantly, a single nucleotide polymorphism (SNP) within *ATP6V1B2*, rs1106634, has exhibited a suggestive p-value in a meta-analysis in schizophrenia and bipolar disorder (p=3.97×10^−6^) and has been associated with lifetime risk of depression (p<0.001) in a recent study (51–53).

In NTR, there were no methylome-wide significant findings. The 10 most significant CpGs (Supplementary Table 12) were annotated to genes implicated in similar processes as those found in GS:SFHS, including the innate immune and psychiatric diseases. Although there were no significant findings, the pattern of results in NTR is similar to significant findings in GS:SFHS.The two CpGs identified in the meta-analysis that were present on both array types were also associated in GS:SFHS and were annotated to genes previously implicated in psychiatric disease and the innate immune system (*SKI, SLC5A10*) (see Table 3). These were not nominally significant in NTR (Supplementary Table 13). cg05603985 was hypomethylated in GS:SFHS and hypermethylated in NTR, and cg27589594 was hypomethylated in both cohorts. Although the signal from one CpG was the same across the two cohorts, further large-scale analyses on DNAm collected on the same array will be needed to uncover the role of antidepressants in relation to psychiatric and immune processes in DNAm.

We were able to show that a MS for self-reported antidepressant use was associated with self-reported antidepressant use, MDD status, and 4 lifestyle factors (BMI, alcohol units, smoking status, and pack years) in a subset of GS:SFHS. In addition, when excluding MDD-associated signals in a second DNAm score (MS-control), we found that the predictor was still associated with antidepressant use, although with reduced variance explained (MS R^2^=2.12%; MS-control R^2^=0.93 %). This suggests that, although antidepressant use may in part be a marker of MDD effects on DNAm, antidepressant prescription associations may be partly independent from the condition for which they are prescribed. However, when including lifestyle factors in the association between MS-control and antidepressant use, the variance explained further decreased (R^2^=0.05%). This may indicate that lifestyle risk factors for MDD may play an important role in this association, even in the absence of an MDD diagnosis.

The decreased variance explained may be in part due to differences in the training sample sizes for the scores (MS_N_=3,799, antidepressant users=585; MS-control_N_=2,791, antidepressant users=195; MS-ns_N_=1,952, antidepressant users=226), although variation explained in lifestyle factors is comparable between the MS and MS-control (MS R^2^: BMI=0.75%; alcohol units=1.93%; smoking status=6.11%; pack years=9.14%; MS-control R^2^: BMI=0.53%; alcohol units=1.56%; smoking status=5.08%; pack years=8.09%).

When smokers were excluded from the training sample, MS-ns associations with all variables remained in the same direction but became non-significant. This suggests that smoking may partially confound the associations between antidepressant use and DNAm. However, when excluding smokers in our MWAS analyses, correlations between the effect sizes in the full-sample and non-smoker MWAS indicated that results were not likely confounded by smoking (r=0.980). Further, effects for all significant CpGs were in the same direction in the reduced sample as in the full sample, and were all nominally significant in the non-smoker sample (Supplementary Table 7). Future studies would benefit from training DNAm predictors of self-reported antidepressant use in larger samples of lifelong non-smokers.Only one CpG overlaps between the CpG sites in our MS and CpG sites comprising an MDD DNAm risk score (MS) in Barbu et al. (2020) (15). This CpG (cg09935388) has been associated with smoking in previous studies (54,55), and may therefore capture smoking-associated effects related to both MDD and antidepressant use. Further, in both studies, MDD MS and antidepressant use MS were both more strongly associated with lifestyle factors than with the trait of interest (MDD MS R^2^ in Barbu et al. (2020): MDD=1.75%, BMI=0.097%, alcohol units=0.7%, smoking status=3.2%, pack years=6.5%; Antidepressant use MS R^2^: antidepressant use=2.12%, BMI=0.75%, alcohol units=1.93%, smoking status=6.11%, pack years=9.14%). The stronger associations for both MS with lifestyle factors may indicate that the scores capture an archive of relevant current and past lifestyle factors that are also associated with depression and the prescription of antidepressants. Previous large-scale studies have found profound effects of each of the lifestyle factors on the methylome (56–58). However, the MS was still associated with antidepressant use after accounting for the lifestyle factors mentioned above (R^2^=0.56%), indicating a possible independent effect of antidepressants. Due to the complex associations between DNAm, MDD, and lifestyle factors, future large-scale studies, including longitudinal studies, are warranted to investigate independent effects of each variable in relation to MDD.

To investigate potential relationships between DNAm links to antidepressant use and MDD, we conducted a MDD MWAS in GS (N=8,844, cases=1,641). While none of the top 10 CpGs in the antidepressant use MWAS were significantly associated with MDD, the effect sizes showed a moderate correlation (r=0.674; See Supplementary Figures 2 and 3). This suggests that DNAm signatures of antidepressant use may partially reflect confounding by indication and much larger samples will be needed to disentangle this relationship.

Using missMethyl, we identified altered DNAm nearby several genes involved in myeloid cell differentiation. Myeloid cells play an important role in the innate immune response, and their activation and differentiation depend in part on epigenetic mechanisms (59). Several of the genes annotated to CpGs identified at genome-wide significance here were associated with the innate immune response in previous GWAS; this may complement the finding that genes annotated to the 144 CpG sites differentially methylated at p<1×10^−5^ are enriched for myeloid cell differentiation.

Further, the differentially-methylated regions were annotated to genes that play a role in cell maintenance, as well as brain and sensory organ development (46,49). Importantly, mutations in GATA2 have previously been related to wide-ranging immunological disorders, providing further evidence linking antidepressant use and the immune system (47).

The role of the innate immune system has previously been investigated in relation to psychiatric disorders. In MDD, studies have found a number of pro-inflammatory cytokines, such as TNF-α and IL-6, to have higher concentrations in depressed individuals, as compared to healthy controls (60). Importantly, antidepressants, particularly SSRIs, have been shown to exert effects on the immune system by causing a reduction in inflammatory markers. Generally, antidepressants have been found to reduce levels of pro-inflammatory cytokine, such as TNF-α, IL-6, and IL-1β, although their effects are complex and incompletely understood (60). The current study suggests novel links between DNAm, antidepressants, and processes involved in the innate immune system. Studies that investigate links between antidepressants and DNAm alterations at CpG sites that are known to be associated with the expression of innate immune system, myeloid cell differentiation, and MDD-specific genes may be of interest.

There are a number of potential limitations to the current study. Firstly, although self-reported antidepressant use could not be restricted to specific dates prior to the blood draw date in GS:SFHS, the variable is considered contemporaneous to blood draw date as self-report questionnaires were sent 1-2 weeks before the clinic appointment. However, the questionnaire did not record types of antidepressants, which may explain the different top CpG sites based on the predictor input in MWAS. The different CpG sites identified in each MWAS may also be due in part to the proportion of overlapping individuals used in each analysis (N_overlap in antidepressant users_=289; N_overlap in controls_=4,811). Nonetheless, as indicated in Supplementary Table 11, most of the CpG sites significantly associated with SSRI use within 12 months prior to blood draw date were nominally significant in the MWAS where self-reported antidepressant use was fit as the predictor. This pattern was also observed when investigating the top 10 CpG sites associated with the self-reported measure in the SSRI use MWAS (Supplementary Table 10). The findings above indicate there is agreement between different forms of data collection, specifically self-reported medication use and record linkage to PIS. The variables have previously been investigated in a study by Hafferty et al. (2018) (61), who showed that self-reported antidepressant use in GS:SFHS showed very good agreement with record linkage data at 3- and 6-month fixed time windows (61).

Secondly, since we chose to restrict PIS data linkage to 12 months before the blood draw date, it seems likely that the DNAm changes reported may occur in response to antidepressant treatment, although this requires further replication. Nevertheless, the DNAm data provided here is cross-sectional, and it is not possible to show how the trajectory of DNAm at each CpG site evolved over time. Future studies should therefore collect DNAm at multiple time points and further address the possibility of confounding by antidepressant indication.

It should also be noted that SSRI prescribing data is unable to confirm whether individuals took the medication as prescribed (62). In addition, the current study is based exclusively on European ancestry cohorts, and the generalisability to diverse ancestries is unknown.

Finally, correlations of effect sizes between the two cohorts are of low magnitude and confidence intervals are large. However, the top 10 CpGs identified in NTR, although not significant, were implicated in similar processes as those in GS:SFHS, and the meta-analysis uncovered two CpGs that were also uncovered in GS:SFHS. These novel findings should serve as a stimulus for larger, international collaborations and meta-analyses in future.

In conclusion, we conducted an MWAS of self-reported antidepressant use across two large, population-based cohorts and identified 10 novel CpG sites in GS:SFHS located in genes previously associated with mental health disorders, implicating the innate immune system. Further, MS predictors were associated with self-reported antidepressant use, MDD, and a number of lifestyle factors in a second sample from the same study. SSRI use derived from linkage health records showed convergent effects indicating agreement with the findings derived using only self-report. Finally, a meta-analysis revealed similar findings to the ones in GS:SFHS. Our findings highlight biological processes that may be relevant to furthering our understanding of antidepressant actions and their side effects.

## Data Availability

Data is available upon request, contacting individuals here: https://www.ed.ac.uk/generation-scotland

## Acknowledgements

Generation Scotland is currently supported by the Wellcome Trust Investigator Award in Science 01/06/2021 to 31/05/26 ‘Exploiting genomic approaches to identify the environmental basis of depression’. (Reference: 220857/Z/20/Z) to McIntosh AM (PI). The DNA methylation profiling and data preparation was supported by Wellcome Investigator Award 220857/Z/20/Z and Grant 104036/Z/14/Z (PI for both grants: McIntosh AM) and through funding from NARSAD (Ref: 27404; PI: Dr DM Howard and Ref: 21956; PI: Dr Kathryn Evans) and the Royal College of Physicians of Edinburgh (Sim Fellowship; PI: Dr HC Whalley). Dr CA was supported by an MRC University Unit award to the MRC Human Genetics Unit, University of Edinburgh, MC_UU_00007/10. Genotyping of the GS:SFHS samples was funded by the MRC and Wellcome Trust [104036/Z/14/Z]. Generation Scotland also receives support from the Chief Scientist Office of the Scottish Government Health Directorates [CZD/16/6] and the Scottish Funding Council [HR03006]. Dr DMH is supported by a Sir Henry Wellcome Postdoctoral Fellowship (Reference 213674/Z/18/Z). Dr MCB is supported by a Guarantors of Brain Non-clinical Post-Doctoral Fellowship. Dr EB is an NIHR Senior Investigator and is a member of the scientific advisory board of Sosei Heptares. Dr ML is supported by a fellowship from the Medical Research Council (MR/S006257/1).

