## Supplementary Materials for "Methylome-wide association study of antidepressant use in Generation Scotland and the Netherlands Twin Register implicates the innate immune system"

**Generation Scotland: further information**

*Selective serotonin reuptake inhibitors (SSRI) prescription and data linkage*

Almost all individuals registered with a General Practitioner (GP) in Scotland are assigned a Community Health Index (CHI) number which acts as a unique identifier. The CHI number was used to record-link GS:SFHS data collected in self-reported questionnaires to the national Prescribing Information System (PIS) administered by NHS National Services Scotland Information Services Division (1). PIS provides information on patient-level prescriptions since April 2009 and allows the identification of medications by approved drug name or paragraph code within the British National Formulary (BNF) (2). GS:SFHS obtained PIS-prescription data for April 2009-March 2011.

*Lifestyle factors and MDD*

Body mass index (BMI) was computed using height (cm) and weight (kg) as measured by clinical staff at baseline recruitment. Alcohol intake was self-reported during the pre-clinical questionnaire, which was administered during the weeks before blood draw date. Participants noted whether they were “never”, “former”, or “current” drinkers; current drinkers were asked: “During the past week, please record how many units of alcohol you have had”. Smoking status was self-reported by participants in response to the question, “Have you ever smoked tobacco?”. Answers were recorded as: “Yes, currently smoke”; “Yes, but stopped within the past 12 months”; “Yes, but stopped more than 12 months ago”; and “No, never smoked”. Pack years for smoking behaviour data were calculated by multiplying the number of cigarette packs (20 cigarettes/pack) smoked per day by the number of years a person has smoked.

Baseline MDD status was assessed by trained researchers who administered an adapted version of the axis-I Structured Clinical Interview of the Diagnostic and Statistical Manual, version IV (SCID) (3). This was administered to participants who answered “Yes” to either of the following screening questions: “Have you ever seen anybody for emotional or psychiatric problems?” and “Was there ever a time when you, or someone else, thought you should see someone because of the way you were feeling or acting?”. Participants with no MDD were defined as those individuals who answered “No” to the two screening questions above and did not fulfil criteria for a current or previous MDD diagnosis following the SCID interview.

*DNA methylation QC*

The raw data for all participants in the present study was pre-processed and quality checked for all individuals, after participant removal due to a number of reasons, including sex mismatch (N_removed_=24), having more than 1% CpG sites with a detection p-value>0.05 (N_removed_=52), showing evidence of dye bias, being an outlier for bisulphite conversion control probes (N_removed_=1), having a median methylated signal intensity more than 3 standard deviations lower than expected (N_removed_=74), and other technical and dataset-related issues (N_removed_=602). A total of 10,495 CpG sites were removed due to low beadcount, poor detection p-value, and sub-optimal binding.

Firstly, R package “minfi” was used to read in the IDAT files, compute M and beta values, and remove probes with large detection p-values, and to compute methylation principal components (PC) of control probes. Secondly, correction was applied for (1) technical variation, where M values were included as outcome variables in a mixed linear model adjusting for appointment date and Sentrix ID (random effects), jointly with Sentrix position, batch, clinic, year, weekday, and 20 methylation PCs (fixed effects); and (2) biological variation by fitting residuals of (1) as outcome variables in a second mixed linear model adjusting for genetic and common family shared environmental contributions (random effects classed as G: common genetic; K: kinship; F: nuclear family; C: couple; and S: sibling) (4) and sex, age, and estimated cell types proportions (CD8T, CD4T, NK, Bcell, Mono, Gran) (fixed effects) (5).

Cross-reactive (N=42,558) and polymorphic (N=10,971) CpGs, obtained from McCartney et al. (2016) were removed from the final dataset, resulting in 674,246 CpGs across the 22 autosomes (6).

*Sensitivity analyses*

Due to the strong impact of smoking on DNAm (7), sensitivity analyses were performed in a subset of individuals that had never smoked (N=3,351, N_antidepressant use_=304). To address confounding by indication for antidepressant use, we conducted MWAS in a subset of antidepressant users defined as (1) individuals who did not have an MDD diagnosis and took antidepressants (N=5,268, antidepressant users=320) and (2) individuals who had an MDD diagnosis and took antidepressants (N=5,368, antidepressant users=420). In both datasets, controls were defined as those who answered no to regularly taking antidepressants.

We also performed the above linear regression using SSRI prescribing data as a predictor, to identify whether there are differences in DNAm in relation to antidepressant use when using SSRI PIS data as a phenotype. We restricted SSRI dispense dates to 12 months prior to the blood draw date for each individual.

*DNAm score analysis*

In wave 1, self-reported antidepressant use (N=3,799; antidepressant users=585) was first residualised for sex, age, and 10 genetic PCs. The R package “biglasso” was then used to train DNAm predictors, where probes were restricted to only those available on the Illumina 450K array (N=365,837), to maximise the availability and use of our predictor to other cohorts using the 450K array. Penalised regression was applied using the “cv.lasso” function and 10-fold cross validation. Non-zero coefficients from this model, with the lambda value corresponding to the mean square error, were then used to create methylation scores for self-reported antidepressant use in GS wave 2.

Due to the known association between smoking and DNAm (7), we also trained DNAm predictors on a set of non-smokers in wave 1, to exclude potentially confounding smoking signals within our predictor. This was achieved by excluding those individuals who had a smoking history (i.e. answered “yes” to the question, “have you ever smoked tobacco?”) from all individuals with self-reported antidepressant use data (N excluded=1,847). The training dataset here consisted of 1,952 individuals (226 antidepressant users). Similarly, self-reported antidepressant use was residualised for age, sex, and 10 genetic PCs, and penalised regression was applied as above.

Finally, to exclude MDD-associated effects within our predictor, we trained a further DNAm score on a set of individuals with no MDD diagnosis in wave 1 (MS-control). The training dataset consisted of 2,791 individuals (antidepressant users=195).

**Netherlands Twin Register: further information**

*MDD*

The Dutch computerized version of the CIDI was administered in a telephone interview in 1997 and 2007 in order to obtain lifetime MDD status according to diagnostic criteria of the DSM-4 (8). The LIDAS was developed as part of the Biobanking and Biomolecular Resources Research Infrastructure (BBMRI-NL) to efficiently identify lifetime MDD in population-based cohorts, and was distributed in 2015-2020 among NTR respondents. LIDAS is based on the Composite International Diagnostic Interview short form (CIDI-sf) (9) and contains diagnostic self-report items to determine lifetime MDD status in accordance with DSM-5 criteria. DSM-oriented scale scores of the Adult Self Report (10) were derived from longitudinal NTR surveys (1991-2013), containing 10 to 14 items with ratings on a 3-point scale. Within-survey scale scores were converted to *z* scores to account for variation in the number of items, and subsequently converted to *T* scores, where a threshold of 69 was applied to identify MDD cases (> 69) and controls (≤ 69).

*DNA methylation QC*

Genomic DNA (500ng) from whole blood was bisulfite treated using the Zymo EZ DNA Methylation kit (Zymo Research Corp, Irvine, CA, USA), and 4 µl of bisulfite-converted DNA was measured on the Illumina 450k array following the manufacturer’s protocol.

A number of sample- and probe-level quality checks and sample identity checks were performed, as described in detail previously (11). In short, sample-level QC was performed using MethylAid (12). Probes were set to missing in a sample if they had an intensity value of exactly zero, or a detection p> .01, or a bead count of<3. After these steps, probes that failed based on the above criteria in >5% of the samples were excluded from all samples (only probes with a success rate ≥ 0.95 were retained). The following probes were also removed: sex chromosomes, probes with a single nucleotide polymorphism (SNP) within the CpG site (at the C or G position) irrespective of minor allele frequency in the Genome of the Netherlands (GoNL) population (13), and ambiguous mapping probes reported by Chen et al. (2013) with an overlap of at least 47 bases per probe (14). The methylation data were normalized with functional normalization in the R package “minfi” (15), a method that removes technical between-sample variation by regressing out PCs estimated from control probes on the methylation array. The first 4 PCs of the internal control probes matrix were significantly correlated with technical variables and together accounted for 89% of technical variation (11). These PCs were added as covariates to the functional normalization function and regressed out. Good quality whole-blood DNAm data were available for 3087 samples from 3055 individuals, including monozygotic and dizygotic twins, parents of twins, siblings of twins and spouses of twins.

**R code**

a. **Methylome-wide association study**: MWAS was performed through a locally-created pipeline; the phenotype input file should contain a column of participant IDs and separate columns for each phenotype; the pipeline also takes as input a covariate file, which should contain a column of participant IDs and separate columns with each covariate. The script below is the main part of the pipeline, indicating the regression model for self-reported antidepressant use MWAS; SSRI use MWAS were performed in a similar fashion.

Input sample: 14832 # initial input file contains 14,832 individuals

Complete phenotype and covariates: 6428 # by merging with DNA methylation data and covariates, there are 6,428 individuals remaining

Probes excluded: 0 # no probes were excluded for this analysis

Model: CpG ~antidep + units + bmi + ever_smoke + pack_years + dep_status + wave

Design matrix: CpG ~ (Intercept) + antidep + units + bmi + ever_smoke + pack_years + dep_status + wave3

MWAS sample size: 6428 # final MWAS sample size

MWAS probes: 674246 # final number of MWAS probes

b. **Manhattan plot:** the manhattan plot function was performed using the script from the following website:

<https://github.com/pcgoddard/Burchardlab_Tutorials/wiki/GGplot2-Manhattan-Plot-Function>

gg.manhattan <- function(df, threshold, hlight, col, ylims, title){

\# format df

df.tmp <- df %>%

\# Compute chromosome size

group_by(CHR) %>%

summarise(chr_len=max(MAPINFO)) %>%

\# Calculate cumulative position of each chromosome

mutate(tot=cumsum(chr_len)-chr_len) %>%

select(-chr_len) %>%

\# Add this info to the initial dataset

left_join(df, ., by=c("CHR"="CHR")) %>%

\# Add a cumulative position of each SNP

arrange(CHR, MAPINFO) %>%

mutate( BPcum=MAPINFO+tot) %>%

\# Add highlight and annotation information

mutate( is_highlight=ifelse(SNP %in% hlight, "yes", "no")) %>%

mutate( is_annotate=ifelse(P < threshold, "yes", "no"))

\# get chromosome center positions for x-axis

axisdf <- df.tmp %>% group_by(CHR) %>% summarize(center=( max(BPcum) + min(BPcum) ) / 2 )

ggplot(df.tmp, aes(x=BPcum, y=-log10(P))) +

\# Show all points

geom_point(aes(color=as.factor(CHR)), alpha=0.8, size=2) +

scale_color_manual(values = rep(col, 22 )) +

\# custom X axis:

scale_x_continuous( label = axisdf$CHR, breaks= axisdf$center ) +

scale_y_continuous(expand = c(0, 0), limits = ylims) + # expand=c(0,0)removes space between plot area and x axis

\# add plot and axis titles

ggtitle(paste0(title)) +

labs(x = "Chromosome") +

\# add genome-wide sig and sugg lines

geom_hline(yintercept = -log10(sig)) +

geom_hline(yintercept = -log10(sugg), linetype="dashed") +

\# Add highlighted points

#geom_point(data=subset(df.tmp, is_highlight=="yes"), color="orange", size=2) +

\# Add label using ggrepel to avoid overlapping

geom_label_repel(data=df.tmp[df.tmp$is_annotate=="yes",], aes(label=as.factor(SNP), alpha=0.7), size=5, force=1.3) +

\# Custom the theme:

theme_bw(base_size = 22) +

theme(

plot.title = element_text(hjust = 0.5),

legend.position="none",

panel.border = element_blank(),

panel.grid.major.x = element_blank(),

panel.grid.minor.x = element_blank()

)

}

\# Variables

mypalette <- c("dark grey", "black", "orangered3") # chr color palette

mysnps <- subset(selfrep_mod3, p_corrected < 0.05)

\# snps to highlight

sig = 7.42e-08 # bonferroni threshold line

sugg = 0.05 # suggestive threshold line

\# Run script on self-rep; 3-month; 6-month; 9-month; and 12-month analyses

\# Change "CHR" to numbers

selfrep_mod3$CHR = sub("chr", "", selfrep_mod3$CHR)

selfrep_mod3$CHR = as.numeric(selfrep_mod3$CHR)

colnames(selfrep_mod3)[1]<-"SNP"

colnames(selfrep_mod3)[10]<-"P"

selfrep_mod3$MAPINFO = as.numeric(selfrep_mod3$MAPINFO)

\# RUN PLOT

tiff("C:/path_to_file/selfrep_manh.tiff",

width=3200, height=2000, res = 200)

gg.manhattan(selfrep_mod3,threshold=7.42e-08, hlight=mysnps, col=mypalette,

ylims=c(0,12), title="Self-reported antidepressant use in GS (N=6,428)")

dev.off()

c. **DNAm risk scores – biglasso and risk score calculation**

**LASSO regression (used for training dataset)**

Install.packages(“glmnet”)

install.packages("methods")

library(glmnet)

library(methods)

beta<- read.table("/path/to/DNAm")

phenotype_file <- read.table("/path/to/phenotype_file” ,header=T)

\# subset methylation data according to whether they have the predictor of interest

pheno <- phenotype_file[!is.na(phenotype_file$pheno),]

a = which(colnames(clean_beta) %in% pheno$ID)

clean_beta1 = clean_beta[,a]

rm(clean_beta)

tclean_beta = t(clean_beta1)

rm(clean_beta1)

ids = rownames(tclean_beta)

dep1 = pheno[match(ids, pheno$ID),]

y = dep1$pheno

\# remove columns with NAs as glmnet can't use missing data

x1 = tclean_beta[,apply(tclean_beta, 2, function(tar) !any(is.na(tar)))]

rm(tclean_beta)

set.seed(1.234)

\# Cross-validation: find the best shrinkage value - the alpha=1 means it's a LASSO model

lasso.cv <- cv.glmnet(x1, y, alpha=1, nfolds=10)

save(lasso.cv, “/lasso/output/")

lambda.min = lasso.cv$lambda.min

m = glmnet(x1,y,lambda=lambda.min)

\# create dataframe with weights for each probe

test = data.frame(coef.name = dimnames(coef(m))[[1]], coef.value = matrix(coef(m)))

\# only take predictors where the co-efficient value is not 0

coef = test[test$coef.value!=0,]

save(coef, “coef/file/")

**DNAm risk score calculation (used for testing dataset)**

```
## Create antidep scores in wave 3 GS
## Read in raw methylation dataset (colnames should be participant IDs; rownames should be CpG sites)
```

data <- read.table("/path/to/DNAm”)

\## Read in list of participants you want to calculate risk scores for (ID column: participant ID; antidepressant use column: 0/1 for non-users/users)

dep_test <- readRDS("/path/to/phenotype_file”)

a = which(colnames(data) %in% dep_test$ID)

meth = data[,a]

rm(data)

dat = meth

rm(meth)

meth = t(dat)

rm(dat)

meth1 = as.data.frame(meth)

meth1$id = as.character(rownames(meth1))

\## Read in LASSO coefficients, which contain coef.name (CpG site); coef.value (CpG weight) --> this was derived in training dataset

load("/path/to/coefficient_file”)

a = which(names(meth1) %in% coef$coef.name)

meth2 = meth1[,a]

meth3 = t(meth2)

probes <- intersect(coef$coef.name, rownames(meth3))

rownames(coef) = coef$coef.name

b = meth3[probes,]

p = coef[probes,]

for (i in probes) {

b[i,]= b[i,]*p[i,"coef.value"]

}

predicted_dep=colSums(b) + coef[1,2]

pred_dep = as.data.frame(predicted_dep)

pred_dep$ID = rownames(pred_dep)

dep = merge(dep_test, pred_dep, by="ID")

\## Save this dataset; this should have: participant ID; antidepressant use phenotype; DNAm risk score as columns

save(dep, “/path/to/risk_score_file”)

| **SSRI dispensing records** | **Prescriptions dispensed in 12 months prior to blood draw date** | **Average time of last dispense date from blood draw** | **Dosage prescribed** | **Drug formulation** |
| --- | --- | --- | --- | --- |
| Citalopram | 1,013 | 80 days | 10mg: 234  20 mg: 592  40 mg: 187 | Tabs: 1,013 |
| Escitalopram | 137 | 76 days | 5 mg: 10  10mg: 87  20 mg: 40 | Tabs: 137 |
| Fluoxetine | 622 | 82 days | 20 mg: 592  60 mg: 25  20 mg/5ml: 5 (solution) | Caps: 617  Solution: 5 |
| Paroxetine | 119 | 63 days | 10mg: 1  20 mg: 94  30 mg: 24 | Tabs: 119 |
| Sertraline | 160 | 78 days | 50 mg: 94  100 mg: 66 | Tabs: 160 |

**Supplementary Table 1.** Descriptive data on 5 SSRIs in N=401 with SSRI prescriptions dispensed in 12 months prior to blood draw date (there were no prescriptions for fluvoxamine maleate); this includes the number of prescriptions in the year before blood draw date; the average time between last dispense date and blood draw date; doses prescribed and drug formulation for each SSRI; 42 participants also have prescriptions for tricyclic and related antidepressant drugs (BNF code: 403010; total prescriptions: 176); and 19 for other antidepressant drugs (BNF code: 403040; total prescriptions: 81).

| **Other medication** | **Self-reported antidepressant use (N=740)** | **No antidepressant use (N=5,688)** |
| --- | --- | --- |
| Cholesterol-lowering medication | 145 (20%) | 532 (9%) |
| Blood pressure-lowering medication | 180 (24%) | 741 (13%) |
| Insulin | 26 (4%) | 76 (1%) |
| Hormone replacement therapy | 60 (8%) | 157 (3%) |
| Oral contraceptive pill | 33 (4%) | 347 (6%) |
| Aspirin | 88 (12%) | 380 (7%) |
| Mood stabilisers | 44 (6%) | 34 (1%) |

**Supplementary Table 2.** Other medications included in the self-report questionnaire. These are: cholesterol-lowering medication; blood pressure–lowering medication; insulin; hormone replacement therapy; oral contraceptive pill or mini pill; Aspirin; and mood stabilizers.

| **Other diagnoses** | **MDD diagnosis (N=1,160)** | **No MDD diagnosis (N=5,268)** |
| --- | --- | --- |
| Heart disease | 33 (3%) | 187 (4%) |
| Stroke | 12 (1%) | 52 (1%) |
| High blood pressure | 181 (16%) | 721 (14%) |
| Diabetes | 46 (4%) | 136 (3%) |
| Alzheimer’s | 0 | 2 (<1%) |
| Parkinson’s | 2 (0.17%) | 7 (<1%) |
| Breast cancer | 18 (2%) | 76 (1%) |
| Bowel cancer | 4 (<1%) | 22 (<1%) |
| Lung cancer | 2 (<1%) | 9 (<1%) |
| Prostate cancer | 2 (<1%) | 17 (<1%) |
| Hip fracture | 5 (<1%) | 30 (<1%) |
| Osteoarthritis | 117 (10%) | 428 (8%) |
| Rheumatoid arthritis | 20 (2%) | 89 (2%) |
| Asthma | 161 (14%) | 590 (11%) |
| COPD | 12 (1%) | 43 (<1%) |

**Supplementary Table 3.** Other diagnoses reported in N=6,428 participants with available self-reported antidepressant use data. The above are the numbers for each disease in those with and without an MDD diagnosis. Some participants have overlapping diagnoses (e.g. heart disease and high blood pressure). There was no overlap with other psychiatric disorders, as these were excluded previously (N=11 with other psychiatric diagnoses, such as bipolar disorder and psychosis).

| **CpG site** | **Beta** |
| --- | --- |
| cg03527086 | -0.004212405 |
| cg07986378 | -0.020272215 |
| cg20379007 | -0.089534147 |
| cg15696038 | 0.021687729 |
| cg19312404 | 0.010117578 |
| cg14215464 | -0.009402831 |
| cg24659858 | 0.002745261 |
| cg17494199 | -0.000933754 |
| cg24162270 | 0.010325688 |
| cg02315513 | 0.010006061 |
| cg24996979 | -0.05186749 |
| cg20858400 | -0.01063382 |
| cg01180441 | -0.003359784 |
| cg04895205 | 0.019848384 |
| cg26923045 | -0.009201063 |
| cg15774391 | -0.006088868 |
| cg13153808 | -0.00206376 |
| cg24174557 | -0.013570738 |
| cg16183741 | -0.010937547 |
| cg02790691 | -0.002922424 |
| cg00530593 | 0.006247074 |
| cg19915582 | 0.001650308 |
| cg11766468 | 0.009234013 |
| cg03636183 | -0.030474026 |
| cg00545759 | -0.000773749 |
| cg01604411 | -0.01739118 |
| cg07626482 | -0.06551365 |
| cg05603985 | -0.112459989 |
| cg16694480 | 0.045996054 |
| cg17465569 | -0.001741683 |
| cg05063806 | -0.009961271 |
| cg04609640 | 0.020506055 |
| cg09935388 | -0.004991975 |
| cg15134919 | -0.010941227 |
| cg19693031 | -0.00497332 |
| cg10117369 | 0.002004 |
| cg07380021 | -0.009811928 |
| cg15743533 | -0.012943329 |
| cg15348679 | 0.008388729 |
| cg09988805 | 0.006476494 |
| cg09902029 | -0.005004094 |
| cg16639998 | -0.006535687 |
| cg06798115 | -0.002743076 |
| cg20496896 | 0.009960566 |
| cg18902742 | -0.001149112 |
| cg17470397 | -0.005131391 |
| ch.4.194519R | -0.02003258 |
| cg16265557 | -0.003987442 |
| cg27324327 | -0.004702437 |
| cg17108430 | -0.013116569 |
| cg27324117 | 0.038234925 |
| cg18304448 | 0.000541316 |
| cg10841124 | 0.00605048 |
| cg08294028 | -0.004832679 |
| cg17508274 | -0.000326059 |
| cg03321829 | -0.003000215 |
| cg20900927 | 0.004271275 |
| cg25798600 | 0.005927247 |
| cg01787834 | -0.007071672 |
| cg06152926 | 0.001857273 |
| cg20160695 | 0.021406791 |
| cg24805875 | -0.007495492 |
| cg23528705 | -0.008169313 |
| cg11165752 | -0.00057982 |
| cg02183564 | 0.005727186 |
| cg07168526 | -0.005590957 |
| cg17440987 | 0.007992899 |
| cg27282922 | -0.001564032 |
| cg17637107 | -0.013347462 |
| cg13566278 | -0.002232548 |
| cg01692968 | -0.007056327 |
| cg13810489 | 0.007524041 |
| cg10310274 | 0.024670046 |
| cg02407308 | 0.002439814 |
| cg13732465 | -0.002694819 |
| cg17443080 | -0.037955157 |

**Supplementary Table 4.** MS CpG sites and weights.

| **CpG site** | **Beta** |
| --- | --- |
| cg06339248 | -0.000943258 |
| cg21709803 | -0.016357319 |
| cg09912988 | -0.00652509 |
| cg02585417 | -0.001346409 |
| cg11152667 | 2.52E-05 |
| cg12778065 | 0.01892422 |
| cg19549714 | -0.000880584 |
| cg19839825 | 0.008791186 |
| cg22797514 | -0.009421663 |
| cg10967587 | -0.015833629 |
| cg20435485 | -0.005177475 |
| cg15796978 | 0.006324823 |
| cg12194939 | -0.005430253 |
| cg06115614 | -0.043073434 |
| cg05781504 | 0.002694968 |
| cg03217966 | -0.000699748 |
| cg25711963 | 0.000407187 |
| cg08647951 | -0.006016662 |
| cg07530264 | 0.044305071 |
| cg13604445 | -0.005637856 |

**Supplementary Table 5.** MS-ns CpG sites and weights.

| **CpG site** | **Beta** |
| --- | --- |
| cg00574958 | -0.00326 |
| cg03713997 | -0.0018 |
| cg24996979 | -0.05164 |
| cg10724690 | -0.02684 |
| cg08022502 | -0.00162 |
| cg02753404 | 0.011948 |
| cg06673794 | 0.013664 |
| cg15443822 | -0.01321 |
| cg25912016 | 0.007045 |
| cg12889746 | -0.01679 |
| cg06653773 | -0.02097 |
| cg13388467 | 0.003218 |
| cg18149561 | -0.0904 |
| cg01387972 | -0.00907 |
| cg03636183 | -0.02859 |
| cg13716638 | -0.00198 |
| cg07626482 | -0.00068 |
| cg24603481 | -0.01121 |
| cg03725309 | -0.03741 |
| cg19693031 | -0.01664 |
| cg05164968 | 0.117529 |
| cg19863073 | -0.00632 |
| cg25119002 | -0.00902 |
| cg03721387 | -0.00546 |
| cg13820699 | -0.00699 |
| cg17887427 | -0.0224 |
| cg04586023 | -0.00459 |
| cg25868769 | -0.00762 |
| cg26337070 | -0.00025 |
| cg16639998 | -0.01056 |
| cg01940273 | -0.02708 |
| cg16265557 | -0.00746 |
| cg05179645 | -0.00099 |
| cg25075684 | 0.008939 |
| cg05423942 | -0.00356 |

**Supplementary Table 6.** MS-control CpG sites and weights.

| **CpG site** | **Full sample MWAS (N=6,428)** | | **Non-smoker MWAS (N=3,351)** | |
| --- | --- | --- | --- | --- |
|  | **β** | **P-value** | **β** | **P-value** |
| cg05603985 | -0.022 | 3.92× 10^−10^ | -0.015 | 0.004 |
| cg05273171 | -0.013 | 2.35× 10^−8^ | -0.013 | 0.001 |
| cg03864397 | -0.030 | 1.05× 10^−8^ | -0.028 | 0.001 |
| cg05186879 | -0.023 | 3.3 × 10^−9^ | -0.017 | 0.006 |
| cg16315329 | -0.039 | 2.52× 10^−8^ | -0.043 | 0.0001 |
| cg25753411 | -0.036 | 2.89× 10^−10^ | -0.036 | 0.0001 |
| cg09511513 | -0.055 | 6.73× 10^−9^ | -0.074 | 1.16x10^-6^ |
| cg26277237 | 0.024 | 2.87× 10^−8^ | 0.032 | 8.45x10^-7^ |
| cg20494891 | -0.026 | 2.32× 10^−8^ | -0.021 | 0.004 |
| cg27589594 | -0.025 | 4.91× 10^−9^ | -0.022 | 0.0008 |

**Supplementary Table 7.** Effect size and nominal p-values for full sample (smokers included) MWAS (N=6,428) and non-smoker MWAS (N=3,351) for CpGs at p < 3.6x10^-8^ in the full-sample MWAS.

| **CpG site** | **Full sample MWAS (N=6,428)** | | **MDD diagnosis + self-reported antidepressant use MWAS (N=5,368)** | | **No MDD diagnosis + self-reported antidepressant use MWAS (N=5,268)** | |
| --- | --- | --- | --- | --- | --- | --- |
|  | **β** | **P-value** | **β** | **P-value** | **β** | **P-value** |
| cg05603985 | -0.022 | 3.92× 10^−10^ | -0.026 | 1.23x10^-8^ | -0.023 | 2.99x10^-6^ |
| cg05273171 | -0.013 | 2.35× 10^−8^ | -0.012 | 1.05x10^-4^ | -0.014 | 9.84x10^-5^ |
| cg03864397 | -0.030 | 1.05× 10^−8^ | -0.027 | 1.52x10^-4^ | -0.03 | 1.21x10^-4^ |
| cg05186879 | -0.023 | 3.3 × 10^−9^ | -0.025 | 4.91x10^-7^ | -0.027 | 7.38x10^-7^ |
| cg16315329 | -0.039 | 2.52× 10^−8^ | -0.029 | 1x10^-3^ | -0.051 | 3.75x10^-7^ |
| cg25753411 | -0.036 | 2.89× 10^−10^ | -0.035 | 1.37x10^-6^ | -0.039 | 1.56x10^-6^ |
| cg09511513 | -0.055 | 6.73× 10^−9^ | -0.055 | 4.95x10^-6^ | -0.056 | 3.72x10^-5^ |
| cg26277237 | 0.024 | 2.87× 10^−8^ | 0.026 | 1.1x10^-6^ | 0.026 | 1.32x10^-5^ |
| cg20494891 | -0.026 | 2.32× 10^−8^ | -0.027 | 5.55x10^-6^ | -0.025 | 1.85x10^-4^ |
| cg27589594 | -0.025 | 4.91× 10^−9^ | -0.027 | 4.99x10^-7^ | -0.03 | 6.55x10^-7^ |

**Supplementary Table 8.** Effect size and nominal p-values for full sample MWAS (N=6,428), MWAS where cases had an MDD diagnosis and also self-reported antidepressant use (N=5,368), and MWAS where cases did not have an MDD diagnosis and self-reported antidepressant use (N=5,268) for CpGs at p < 3.6x10^-8^ in the full sample MWAS.

| **12-month interval** | | | | | | | |
| --- | --- | --- | --- | --- | --- | --- | --- |
| **CpG site** | **Gene** | **Chrom** | **β** | **P-value** | **P-corr** | **CpG site information** | **Gene information** |
| cg00477015 | *-* | 1 | -0.010 | 4.61× 10^−8^ | 0.033 | - | - |
| cg05954120 | *TMEM79* | 1 | -0.010 | 4.93× 10^−8^ | 0.0352 | HIV infection, gestational age, human liver in subjects with type 2 diabetes, lipid-related phenotypes | Hemoglobin measurement; mean corpuscular haemoglobin concentration; Contributes to epidermal integrity and skin barrier function |
| cg27277419 | *UBR4* | 1 | -0.011 | 4.36× 10^−8^ | 0.031 | - | Immunoglobulin isotype switching measurement; related pathways: innate immune system, MHC mediated antigen processing and presentation |
| cg02652457 | *NR2C2* | 3 | -0.029 | 3.42× 10^−8^ | 0.024 | - | Diseases: premature aging, lateral myocardial infarction; related pathways: gene expression, pathogenesis of cardiovascular disease |
| cg24182240 | *-* | 6 | -0.023 | 1.00× 10^−8^ | 0.007 | - | - |
| cg03924915 | *TRIM38* | 6 | -0.06 | 3.05× 10^−9^ | 0.002 | - | Well-being; height; Related pathways: innate immune system; interferon gamma signalling |
| cg22528063 | *LOC100129034* | 9 | -0.017 | 2.15× 10^−8^ | 0.015 | - | - |
| cg20549160 | *PML* | 15 | -0.012 | 1.92× 10^−8^ | 0.0137 | - | Height; erythrocyte and leukocyte count; insomnia measurement |
| cg04582760 | *UNC45A* | 15 | -0.020 | 3.27× 10^−8^ | 0.023 | - | Regulatory component of progesterone receptor; essential for normal cell proliferation and accumulation of myosin during muscle cell development |
| cg08227615 | *GNB5* | 15 | -0.013 | 6.82× 10^−8^ | 0.049 | - | Intelligence; uveal melanoma; Diseases: intellectual developmental disorder with cardiac arrhythmia; language delay and ADHD; cognitive impairment |
| cg06356163 | *ARHGAP17* | 16 | -0.051 | 3.76× 10^−8^ | 0.027 | - | Appendicular lean mass; blood, urinary, metabolite measurement; chronic kidney disease |

**Supplementary Table 9.** CpG sites significantly associated with dispensed SSRI prescriptions within 12 months prior to the blood draw date (N=12 CpG sites), along with gene annotations, chromosome, standardised effect size, nominal and multiple comparison-corrected p-values. Background information for each CpG site and gene was extracted from EWAS (<http://www.ewascatalog.org/>) and GWAS (<https://www.ebi.ac.uk/gwas/>) catalogue databases.

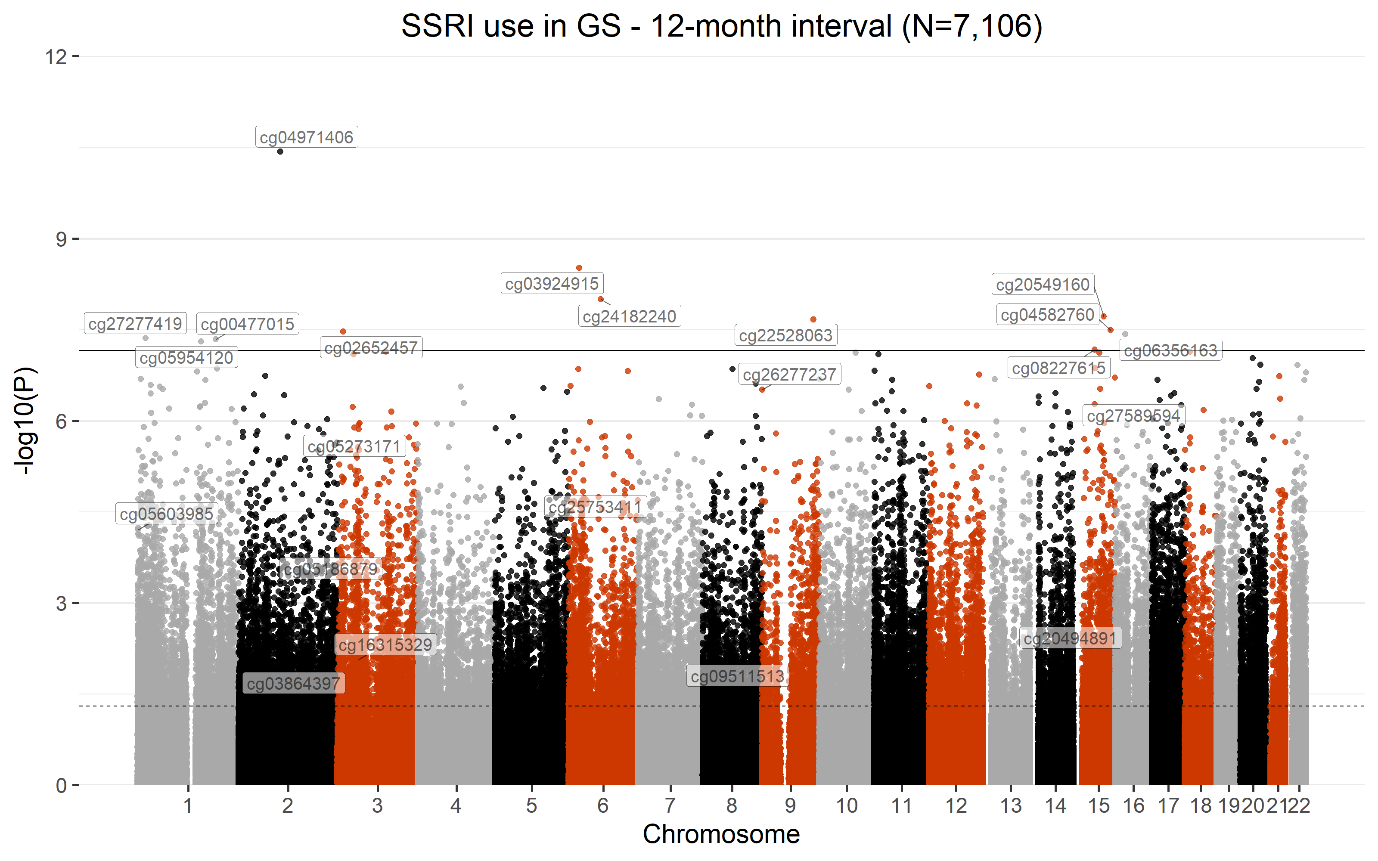

**Supplementary Figure 1.** Manhattan plot showing MWAS of dispensed SSRI prescriptions within 12 months prior to blood draw date. The black line defines the threshold for methylome-wide significance (p≤7.42 x 10^-8^) and the dotted line defines the threshold for nominal significance (p≤0.05). CpG sites associated with dispensed SSRI prescriptions within this time interval are indicated above the methylome-wide significance line. The 10 CpG sites shown to be associated with self-reported antidepressant use are indicated on the plot, between the nominal and methylome-wide significance lines.

| **MWAS** | **CpG site** | **Gene** | **Chrom** | **β** | **P-value** |
| --- | --- | --- | --- | --- | --- |
| **12-month interval** | cg03864397 | CASP10 | 2 | -0.017 | 0.011 |
|  | cg05186879 | MAPKAPK3; CISH | 3 | -0.019 | 0.0001 |
|  | cg05273171 | PNKD; TMBIM1 | 2 | -0.013 | 4.62 x 10^-6^ |
|  | cg05603985 | SKI | 1 | -0.018 | 6.23 x 10^-5^ |
|  | cg09511513 | ATP6V1B2 | 8 | -0.026 | 0.028 |
|  | cg16315329 | MAPKAPK3; CISH | 3 | -0.023 | 0.009 |
|  | cg20494891 | MYO1E | 15 | -0.018 | 0.002 |
|  | cg25753411 | HEBP2 | 6 | -0.031 | 1.41 x 10^-5^ |
|  | cg26277237 | KANK1 | 9 | 0.027 | 3.09 x 10^-7^ |
|  | cg27589594 | SLC5A10 | 17 | -0.027 | 4.56 x 10^-7^ |

**Supplementary Table 10.** Results for CpG sites significantly associated with self-reported antidepressant use in the MWAS using dispensed SSRI prescriptions within 12 months prior to the blood draw date. The table incudes gene annotations, chromosome, standardised effect size, and p-value (all p-values are nominally significant).

| **MWAS** | **CpG site** | **Gene** | **Chrom** | **β** | **P-value** |
| --- | --- | --- | --- | --- | --- |
| **Self-reported AD use: 12-month interval** | cg00477015 | - | 1 | 0.001 | 0.6* |
|  | cg02652457 | NR2C2 | 3 | -0.008 | 0.046 |
|  | cg03924915 | TRIM38 | 6 | -0.023 | 0.004 |
|  | cg04582760 | UNC45A | 15 | -0.008 | 0.004 |
|  | cg04971406 | - | 2 | 0.002 | 0.026 |
|  | cg05954120 | TMEM79 | 1 | -0.004 | 0.005 |
|  | cg06356163 | ARHGAP17 | 16 | -0.035 | 1.76 x 10^-6^ |
|  | cg08227615 | GNB5 | 15 | -0.009 | 2.82 x 10^-6^ |
|  | cg20549160 | PML | 15 | -0.008 | 1.03 x 10^-5^ |
|  | cg22528063 | LOC100129034 | 9 | -0.006 | 0.01 |
|  | cg24182240 | - | 6 | 0.003 | 0.292* |
|  | cg27277419 | UBR4 | 1 | -0.005 | 0.002 |

**Supplementary Table 11.** Results for CpG sites significantly associated with dispensed SSRI prescriptions within 12 months prior to the blood draw date in the MWAS fitting self-reported antidepressant use as predictor; AD: antidepressant. The table incudes gene annotations, chromosome, standardised effect size, and p-value (all p-values are nominally significant). *=indicates associations that are not nominally significant for self-reported antidepressant use.

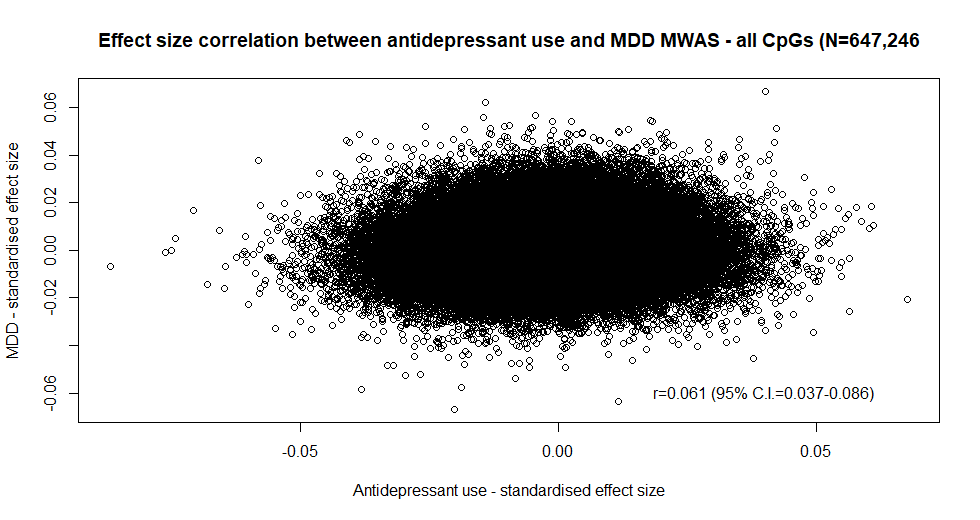

**Supplementary Figure 2.** Correlation plot between antidepressant use MWAS (N=6,428) and MDD MWAS (N=8,419) – all CpGs (N=674,246); *r*=0.061 (95% C.I.=0.037-0.086).

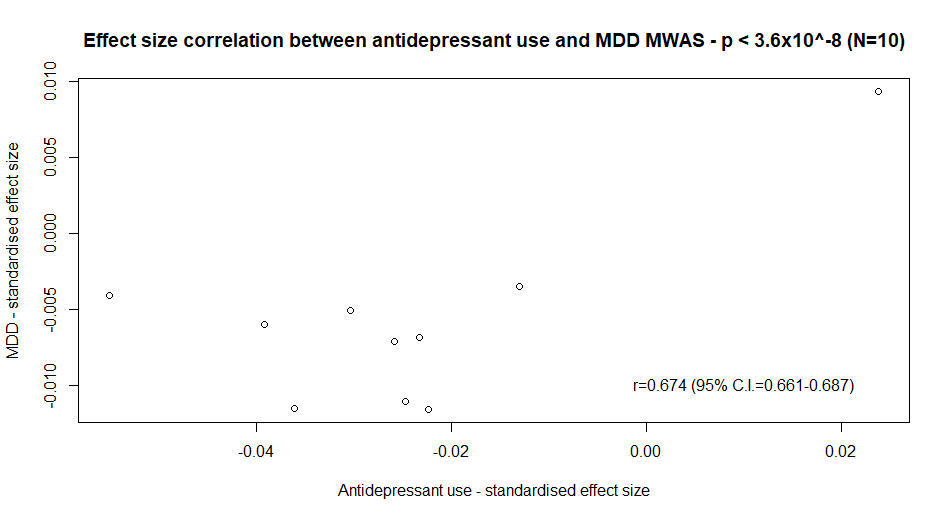

**Supplementary Figure 3.** Correlation plot between antidepressant use MWAS (N=6,428) and MDD MWAS (N=8,419) - CpGs at p < 3.6x10^-8^ (N=10); *r*=0.674 (95% C.I.=0.661-0.687).

| **CpG site** | **Gene** | **Chrom** | **ß** | **P-value** | **CpG site information** | **Gene information** |
| --- | --- | --- | --- | --- | --- | --- |
| cg26168975 | C1orf58 | 1 | 0.255 | 5.36E-07 | - | - |
| cg10333415 | C3orf21 | 3 | 0.104 | 6.47E-07 | - | - |
| cg11307865 | - | 12 | 0.13 | 9.43E-07 | Age from birth to adolescence (16), age at menarche (17) | - |
| cg11750706 | PRDM2 | 1 | 0.133 | 1.16E-06 | Age from birth to adolescence (16) | Age at menarche (18), brain aneurysm (19), male puberty onset (20), PHF-tau measurement (21) |
| cg14256102 | ZNF615 | 19 | -0.125 | 1.30E-06 | - | - |
| cg23514110 | CFD | 19 | -0.224 | 1.32E-06 | Age from birth to adolescence (16) | Reticulocyte count (22), neurofibrillary tangles (21) |
| cg26268742 | PLA2G4C | 19 | 0.128 | 1.47E-06 | Age from birth to adolescence (16) | Acute myeloid leukaemia (23) |
| cg02798801 | BUD31 | 7 | 0.152 | 1.55E-06 | Age from birth to adolescence (16) | Risk-taking behaviour (24,25) |
| cg04985396 | NEGR1 | 1 | -0.122 | 2.56E-06 | Age from birth to adolescence (16,26), HIV infection (27) | BMI (28–30), unipolar depression (31), educational attainment (32), cognitive function measurement (32), mathematical ability (32), age at menarche (18), intelligence (33), depressive symptoms (34) |
| cg01709316 | KCTD5 | 16 | 0.331 | 3.25E-06 | Age from birth to adolescence (16), Rheumatoid arthritis (35) | Body fat (18), neurofibrillary tangles (21) |

**Supplementary Table 12.** Results for top 10 CpG sites in NTR (N=2,449, antidepressant use=74) along with gene annotations, chromosome, standardised effect size, nominal and Bonferroni-corrected p-values. Background information for each CpG and gene was extracted from EWAS (<http://www.ewascatalog.org/>; association between traits and CpGs on Illumina 450K array at p≤1.0x10^-4)^; and GWAS (<https://www.ebi.ac.uk/gwas/>; associations between traits and SNPs at p≤1.0x10^-5^) catalogue databases. All associations included in the table from these two catalogues are genome-wide significant.

**Concordant signals between GS and NTR**

There were N=318,856 overlapping CpGs annotated to 242,529 genes between GS:SFHS and NTR, of which N=71,823 were hypermethylated (annotated to N=52,392 genes) and N=92,351 were hypomethylated (annotated to N=71,735 genes) in both cohorts.

The correlation between effect sizes in the hypermethylated subset was *r*=0.038 (95% C.I.=0.031-0.045), and in the hypomethylated subset was *r*=0.098 (95% C.I.=0.0.92-0.105). In both subsets, the largest number of CpGs were on chromosome 1 (N=7,279 in the hypermethylated subset and N=8,769 in the hypomethylated subset). In the hypermethylated subset most CpGs were localised in open sea area (regions of the genome that are not in the CpG-rich islands and that are subject to greater individual variability (36), N=33,186) and in the hypomethylated subset most CpGs were localised in the island area (regions of the genome containing a large number of CpGs, N=33,950). Previous evidence shows that the majority of CpG islands are hypomethylated, which is consistent with results here (37).

| **Cohort** | **CpG site** | **Gene** | **Chrom** | **β** | **P-value** |
| --- | --- | --- | --- | --- | --- |
| **GS:SFHS significant CpGs in the NTR MWAS** | cg05603985 | SKI | 1 | 0.024 | 0.189 |
|  | cg27589594 | SLC5A10 | 17 | -0.004 | 0.775 |

**Supplementary Table 13.** CpG sites significantly associated with self-reported antidepressant use in GS:SFHS in the NTR MWAS (N=2). cg05603985 was hypomethylated in GS:SFHS and hypermethylated in NTR, and cg27589594 was hypomethylated in both cohorts.
